# Menopause Hormone Replacement Therapy and Lifestyle Factors affect Metabolism and Immune System in the Serum Proteome of Aging Individuals

**DOI:** 10.1101/2024.06.22.24309293

**Authors:** Clemens Dierks, Roza Sürme Mizrak, Orr Shomroni, Vadim Farztdinov, Kathrin Textoris-Taube, Daniela Ludwig, Johannes Rainer, Michael Mülleder, Ilja Demuth, Markus Ralser

## Abstract

Aging is a fundamental risk factor for a wide array of diseases. The Berlin Aging Study II (BASE-II) is a cohort study designed to investigate the physical, mental, and social determinants of successful aging. We utilized high-throughput mass spectrometry to measure the proteomes of 1890 BASE-II participants, divided into two age groups: 27-37 years and 60-85 years. We employed multiple linear regression analyses to explore the effects of demographic factors such as age, sex, and BMI, along with hormonal treatments and lifestyle factors, on the serum proteome. We identify new associations and confirm previously described proteins linked to age, sex, BMI and hormonal contraceptive use (HCU). Notably, we observed that the abundance of nutrient transport proteins, particularly apolipoproteins, is linked to metabolic diseases in aged individuals, including metabolic syndrome and type 2 diabetes. Additionally, we identified specific alterations explained by lifestyle factors, such as smoking and alcohol consumption. We further report a significant proteome signature in female study participants corresponding to menopause hormone replacement therapy (MHT). We successfully classified these participants based on MHT status with an AUROC of 0.82 using two proteins, Complement Component 9 and Plasminogen, slightly outperforming estradiol (AUROC: 0.80), the active ingredient in most MHT preparations. Overall, our study underscores the impact of lifestyle and hormonal therapies on the serum proteome during aging, primarily affecting components of the immune system and metabolism.

## Introduction

Aging is a primary risk factor for numerous chronic diseases and is associated with health deterioration and functional decline of the body, making it particularly important to investigate molecular changes that are related to age ^1^. While the underlying mechanisms are complex, the serum proteome offers huge potential for deciphering the interrelations of health and disease ^2,3^. The serum proteome responds to changes underlying aging, such as increasing levels of inflammatory proteins and decreasing tissue repair and cell regeneration activity ^4^. Circulating serum protein level levels can help to differentiate between healthy aging and pathological conditions ^1,5–7^. Moreover, an increased understanding of the relationship between chronological and biological aging can contribute to delay or prevent diseases where chronological age is a major risk factor ^6^ as well as improving patient stratification ^1^. Identifying markers of healthy aging is crucial to define appropriate lifestyle interventions and thereby enhancing lifespan ^8^.

Broadly speaking, the serum contains two sets of proteins. Proteins that function in the blood, such as nutrient transport proteins, signaling molecules, proteins of the immune system and the coagulation machinery, and proteins, who circulate in the blood due to tissue leakage, cell necrosis, or to their presence in secreted extracellular vesicles. Several, but not all, of the biomarkers that are currently in use, are part of the former, and are often found within the highly abundant fraction of the plasma proteome, that is easily accessible and simplifies the development of routine applicable biomarker assays ^9–12^. In recent years, the application of multiplexed proteome technologies has enhanced our ability to probe the complex molecular landscape of the proteome in plasma or serum. Recent advancements in mass spectrometry-based proteomic platforms have addressed previous challenges related to throughput and cost, enabling large-scale recording of plasma proteomes ^13^.

In this study, we recorded the serum proteomes of 1890 participants from the Berlin Aging Study II (BASE-II)^14^ The study aims to uncover factors contributing to healthy aging, involving in total 2200, with about two thirds belonging to the older group aged 60 to 85 years and about one third representing the younger group of participants aged between 27 to 37 years from the metropolitan area of Berlin, Germany. We obtained a quantitative dataset focussing on the neat serum proteome and the high abundant fraction of the serum proteome, and quantifies acute phase proteins, lipid transport proteins, proteins involved in the innate or adaptive immune system and coagulation factors. We report their dependency on age, common disease, selected lifestyle factors, and discover strong signals that are attributable to hormonal replacement therapy.

## Results

### Study Profile and Proteomic Assessment of the Berlin Aging Study II

We conducted MS-based proteomic measurements on a total of 2091 serum samples, including 105 serum control and 96 pooled study samples, to generate a proteomic baseline from the BASE-II. The samples analyzed are derived from two age groups: older adults (N = 1433) aged 60 to 85 years and younger adults (N = 426) aged 27 to 37 years. Study demographics are given in Table 1.

**Table 1:**
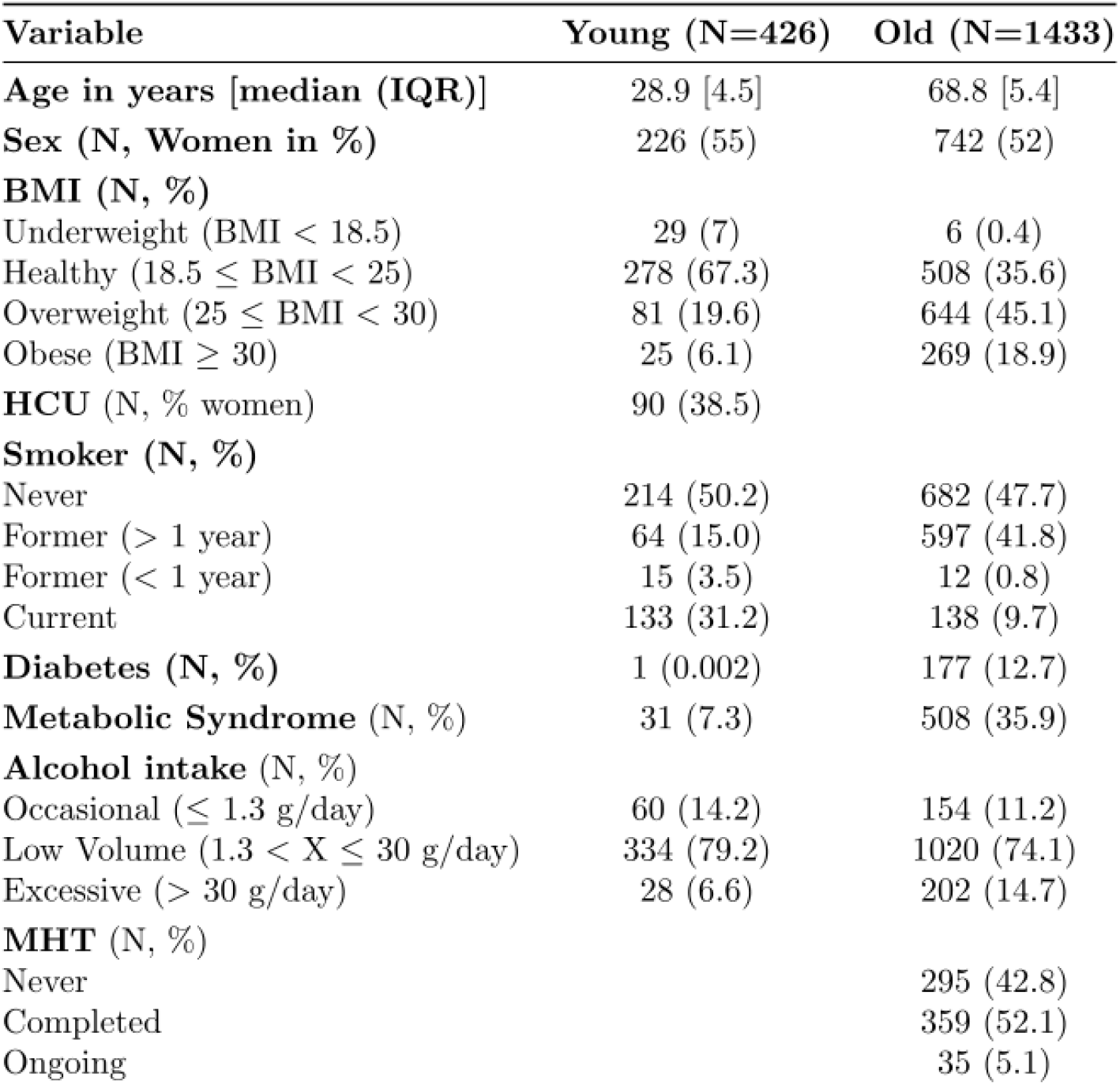
Study Demographics of study participants subjected to the analysis stratified by age groups. HCU: Hormonal Contraceptive Use, MHT: Menopause Hormone Replacement Therapy.

The samples were prepared for measurement in a semi-automatic workflow that allows for high-throughput and consistent sample quality including tryptic digestion and sample cleanup with solid-phase extraction ^15^. Samples were divided into 8 batches, each consisting of three 96-well plates, and were subjected to mass spectrometric measurement using a timsTOF Pro1 mass analyzer (Bruker Daltonics) operating in diaPASEF mode, using analytical flow rate chromatography using ^16,17^. Raw MS data was processed by DIA-NN ^18,19^, using the DiOGenes spectral library ^20^. In the subsequent preprocessing the data was normalized, filtered for outlier samples and low-presence peptides, imputed and corrected for plate effects (N = 24). Among the data set, 31 samples were removed due to low technical quality, or missing metadata (age and BMI) leaving the dataset with a total of 1859 proteomic samples from BASE-II participants. After this preprocessing, the final dataset quantified 2079 proteotypic peptides derived from 248 protein groups (Table S1).

We calculated a median coefficient of variation (CV) of protein quantities of 12.74 % in study pools and 10.89 % for serum controls samples (Zen-Bio). Reflecting the biological signal, protein quantities in study samples deviated from the technical controls (CV of 27.79 %; Table S1)). While most of the proteins are normally distributed, some proteins, such as lipoprotein (a) (Lp(a)) and complement factor H-related protein 1 (CFHR1), exhibited tailed or almost bimodal distributions (Figures S59, S158). Tailed distributions in certain proteins, particularly pronounced among the younger age group, were found for proteins that we recently associated to the intake of hormonal contraceptives in younger women (including angiotensinogen (AGT), fetuin B (FETUB), sex hormone binding globulin (SHBG), plasminogen (PLG) or peptidoglycan recognition protein 2 (PGLYRP2))^21^. (Figures S2 - S254 for Log2 transformed protein abundance distributions of all proteins across the entire cohort, as well as stratified by age groups).

We analyzed functional groups, such as immunoglobulins, coagulation factors, acute-phase proteins, complement factors and lipid transport proteins for their variation. Immunoglobulins (N = 60 protein groups) had the highest median relative variation (CV 44.3 %, range: 23.6 - 264.8 %), and included the proteins with the highest individual relative CVs such as IGHV3-11 (264.8 %), IGHV3-21 (183.3 %) and IGLV4-60 (150.6 %). In contrast, proteins involved in the complement system (N = 36) exhibited lower variation (median CV: 18.4 %, range: 10 - 63.5 %), including some of the least variable proteins such as the two central mediators of the component system C5 and C3 (both 10.4%). Proteins related to coagulation (N = 30), lipid transport (N = 20) and acute-phase response (N = 26), showed CVs of 19.6 % (range: 11.5% - 138.2%), 23.5% (12.9 % −102%) and 22% (10.4% - 262.6%). Besides IGHV3-11, serum amyloid A1 (SAA1; 262.6%) and the pregnancy zone protein (PZP, 225.1%) showed the highest variation in the study samples, while hemopexin (HPX, 9.2%), complement factor 1R (C1R, 10%) and attractin (ATRN, 10.2%) the lowest. CVs for study samples, pooled study samples and QC samples are shown in Tables S3-S5. Furthermore, a detailed functional analysis of the proteins in the dataset is provided in Table S2.

Next, we conducted a correlation analysis between the values obtained with routinely applied biomarker assays and the corresponding protein abundance levels. For interpreting these results, it is important to keep in mind that proteomics and the diagnostic tests are typically targeting related but not identical analytes. We obtained significant correlations, displayed as spearman’s rho if not described otherwise, between proteomics and biomarker assays for apolipoprotein A1 levels (0.67) and B (0.72), low density lipoproteins (LDL) with APOB (0.72) and high-density lipoproteins (HDL) with APOA1 (0.66). Triglyceride measurements showed a moderate correlation with APOC3 levels (0.44). In contrast, diagnostic assays and protein abundance determined by proteomics correlated weekly for albumin (0.32), hemoglobin with the MS acquired hemoglobin subunit alpha 2 (HBA2; 0.31), and for fibrinogen compared to fibrinogen alpha chain levels (0.15). Notably, routine test sex hormone binding globulin (SHBG) measurements (prior to log2 transformation) displayed a robust, albeit not strictly linear relationship, with proteomic data (Kendall’s tau: 0.61, see Figure S250h). The diverging results show that proteomic measurements provide complementary data to establish diagnostic assays (see Figure S250).

### Hormonal contraceptive usage (HCU) is a dominating covariate affecting the serum proteome in young women

We investigated the effect of demographic covariates, such as sex, age, BMI, and HCU due to previous findings ^21^ on the serum proteome of the 1890 BASE-II participants of both age groups by fitting multiple linear regression models to the protein abundance data. In our analysis BMI was treated as a categorical variable comparing underweight (U, BMI < 18.5), overweight (Or, 25 < BMI ≤ 30) and obese (Ob, BMI > 30) participants to those with a healthy BMI (18.5 < BMI ≤ 25). This categorization was chosen because these groups have higher clinical relevance than the actual, numerical BMI value and are commonly used in clinical studies. Besides the significance level of 0.05, we also required the abundance difference of the protein to be higher than the CV of the pooled study samples. Statistical summaries of the association analysis for the entire cohort and separated by age groups are reported inTables S3-S5.

We found 106 protein groups associated with age. Of these, 65 have also been identified as age-dependent in orthogonal studies ^21–30^. Among the 41 new associations identified, 32 were immunoglobulin chains. Other newly found age-associated proteins include serum amyloid A4 (SAA4), proteoglycan 4 and the two platelet-related serum proteins, platelet factor 4 variant 1 (PF4V1) and pro-platelet basic protein (PPBP). Amongst the group of age-associated proteins, we report the strongest negative association, with up to 20% abundance change in 10 years, for serpin family F member 2 (SERPINF2), immunoglobulin delta heavy chain (P0DOX3) and transferrin (TF). Highest increase with progressing age was found in complement component 5 (C5), apolipoprotein B (APOB) and lumican (LUM).

The above defined age-related differences in the proteome were in part sex dependent, with 23 proteins showing a different age-response in male and female study participants. We detected the highest significant differences between men and women in the pregnancy zone protein (PZP), glycosylphosphatidylinositol specific phospholipase D1 (GPLD1), complement component 6 (C6), adiponectin (ADIPOQ) and ceruloplasmin (CP). None of the proteins investigated were associated with an underweight BMI, however, 19 proteins were linked to overweight and obesity, respectively, with APOC2 and fibulin 1 (FBLN1) showing novel associations with elevated BMI ^21,43–47^.

We found new associations with HCU in 15 out of 55 proteins that had not been reported in the related studies before ^21,48–50^, such as the phospholipid transfer protein (PLTP), monocyte differentiation antigen (CD14), alpha-1-antichymotrypsin (SERPINA3) and the coagulation factor V (F5) (see Figure 2). Among all covariates, highest effect sizes and significance were reported for HCU (see Figure 1D) with the top 5 candidates being angiotensinogen (AGT), peptidoglycan recognition protein 2 (PGLYRP2), serpin family A member 6 (SERPINA6), fetuin B (FETUB) and ceruloplasmin (CP). We recently investigated the plasma proteomes of 3,472 participants from the CHRIS cohort ^21,51^, an Alpine population study examining the genetic and molecular backgrounds influencing the incidence and progression of common chronic diseases. To validate our previous findings and demonstrate reproducibility, we compared the effect sizes for age, sex, and BMI categories for the overlapping proteome from the BASE-II analysis with those obtained from a similarly conducted association analysis of the CHRIS dataset, both measured on similar mass spectrometry platforms. We observed high concordance: age (Spearman’s rho: 0.69), sex (0.85), BMI (obese category: 0.73), and HCU (0.9) (see Figures S256). However, the effect sizes for the underweight BMI category showed a low correlation between the studies (−0.25). Among the 125 overlapping proteins identified in both proteomic datasets, we found 53 proteins to be age-dependent and 13 proteins to be sex-dependent in both studies. The age-associated proteins include various functional groups involved in disease pathology, such as components of the complement cascade (C5, C7, C9, C4A, C4BPA, C1, CFD, CFI), apolipoproteins (APOB, APOC3, APOD, APOE, APOH), and proteins involved in blood clotting, including coagulation factors X (F10) and XII (F12) and antithrombin III (SERPINC1). Additionally, we identified acute-phase proteins such as haptoglobin (HP) and transthyretin (TTR).

**Figure 1:**
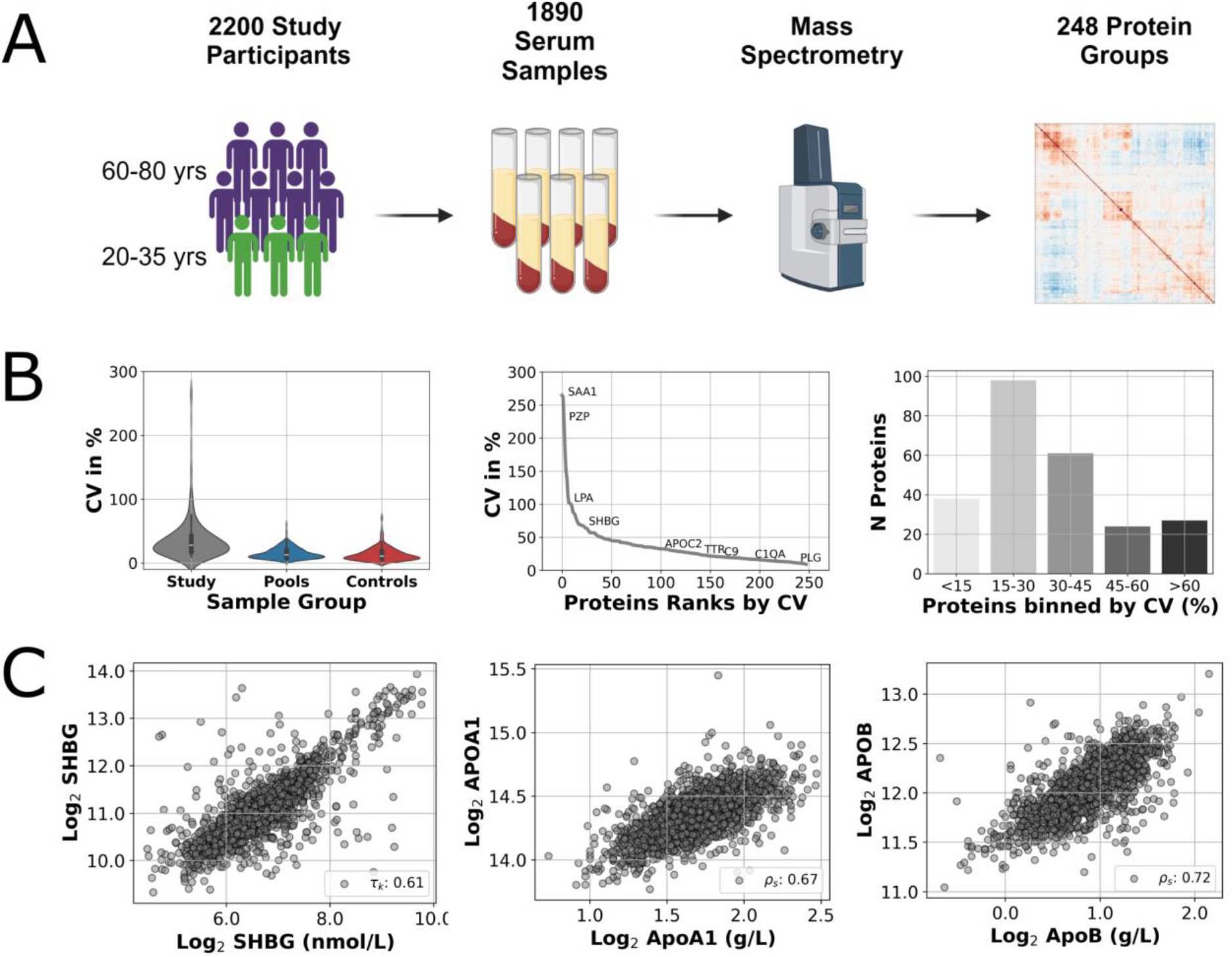
(A) Study Design. Schematic representation of the 2200 BASE-II participants, divided into an older and a younger control group. Serum samples of 1890 participants were analyzed using a high-throughput proteomics platform combining semi-automated sample preparation and latest trapped ion mobility spectrometry. (B) Distribution of coefficient of variation (CV) per protein for study data (gray), pooled study samples (blue) and standardized control serum samples (red) (Violin plots). The line plot depicts proteins ranked by their CV, while barplots show the CV distribution across all proteins. (C) Correlations of sex hormone binding globulin (SHBG), apolipoprotein A1 (APOA1) and B (APOB) concentrations on the y-axis versus the values obtained with related or corresponding routine diagnostic tests (left to right). Diagnostic test values were transformed to log2 for visualization purposes.

**Figure 2:**
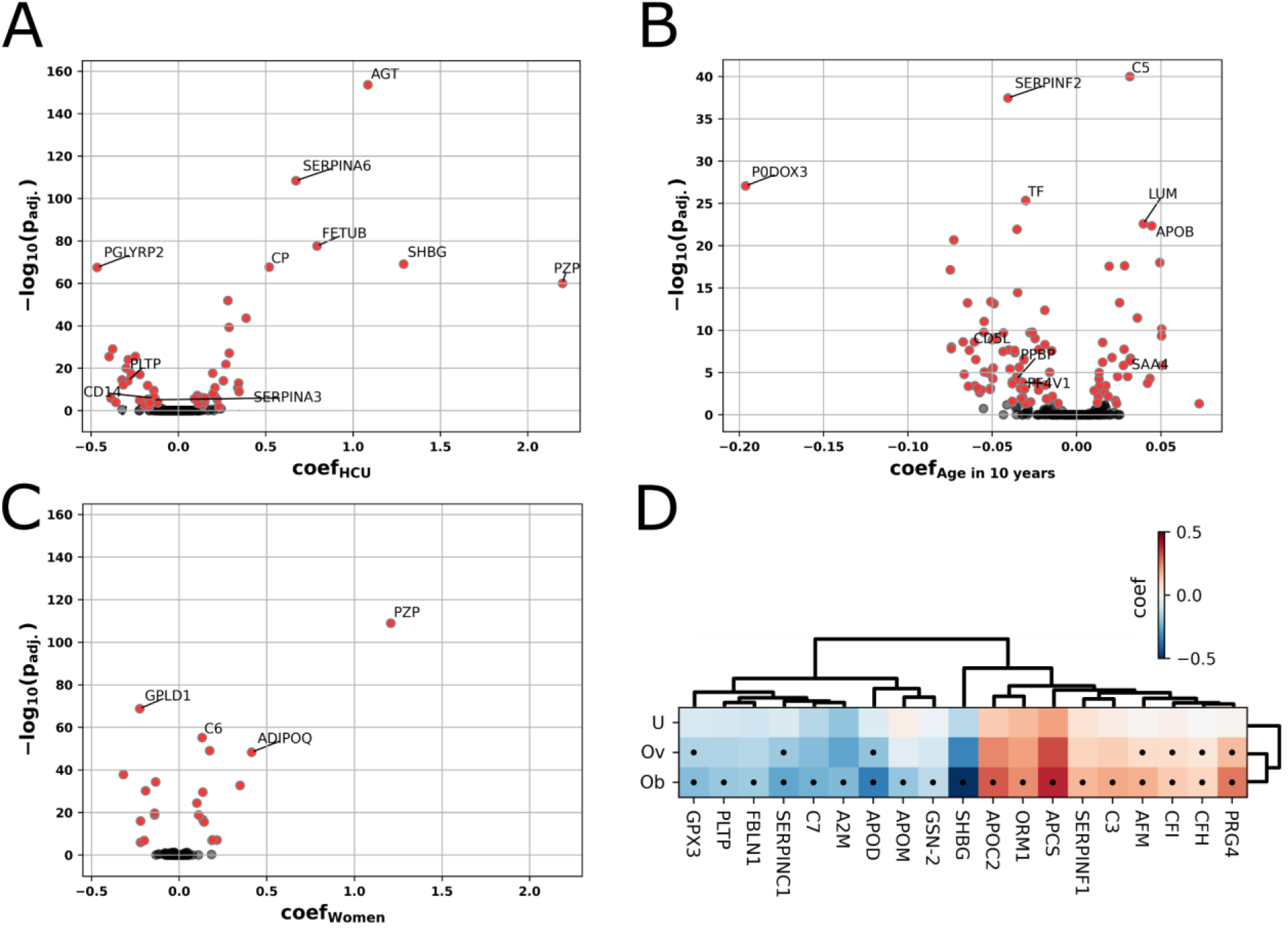
Multiple linear regression was employed to identify significant protein associations with hormonal contraceptive use (HCU), age, sex and different BMI categories (U: Underweight, Ov: Overweight, Ob: Obese). (A) The impact of HCU on the serum proteome. Proteins with largest coefficients and highest significance (in negative log10 adjusted p-values) for proteins with use of hormonal contraceptives. (B) The impact of 10-years of aging on the serum proteome. (C) The impact of sex (men vs. women) on the plasma proteome in Base-II. Proteins of significance are highlighted in red, and gene names are annotated for those discussed in the text. (D) Proteins associated with the different BMI categories (heatmap). Black dots denote significant associations after multiple testing corrections, while color indicates effect size by its coefficient.

### Lifestyle factors show strong associations with apolipoproteins and inflammation markers in the serum proteome

Building on existing knowledge of lifestyle factors impacting the serum proteome^52^, we investigated the effects of smoking habits, alcohol consumption, diabetes mellitus type 2 (T2DM), or metabolic syndrome (metS) on protein abundance as covariates in our multiple regression models in addition to age, sex, BMI and HCU. We only included HCU as a covariate in the younger age group. Adjusted p-values, coefficients and effect sizes obtained from the regression analyses of significantly associated proteins with lifestyle factors and comorbidities are reported in Tables S8 - S12. On this basis we addressed the association of selected individual lifestyle factors, i.e. smoking or alcohol consumption, with the serum proteome.

First, we addressed proteomic changes in current and former smokers. No significantly regulated proteins were found in former smokers that quit smoking more than a year ago. On the other hand, older participants who recently quit smoking (less than a year) had a significantly elevated level of polymeric immunoglobulin receptor (PIGR), which was also true for current smokers compared to participants who had never smoked (n = 896, see Figure 3B). Additionally, immunoglobulin heavy constant gamma 2 (IGHG2) levels were significantly decreased in current smokers from the older age group. IGHG2 levels were also found lowered in the younger group, albeit not significant. Alcohol consumption of the participants was grouped into occasional (< 1.3 g/day), low volume and excessive drinkers (>30 g/day) based on their self-reported habits. Among the 8 significantly associated proteins, the most changes were obtained for apolipoprotein C3 (APOC3) and thyroxine-binding globulin (SERPINA7) (see Figure 3C). Both proteins were significantly changed in low volume and excessive drinkers compared to participants that only occasionally drink alcohol, though significance increased with age.

**Figure 3:**
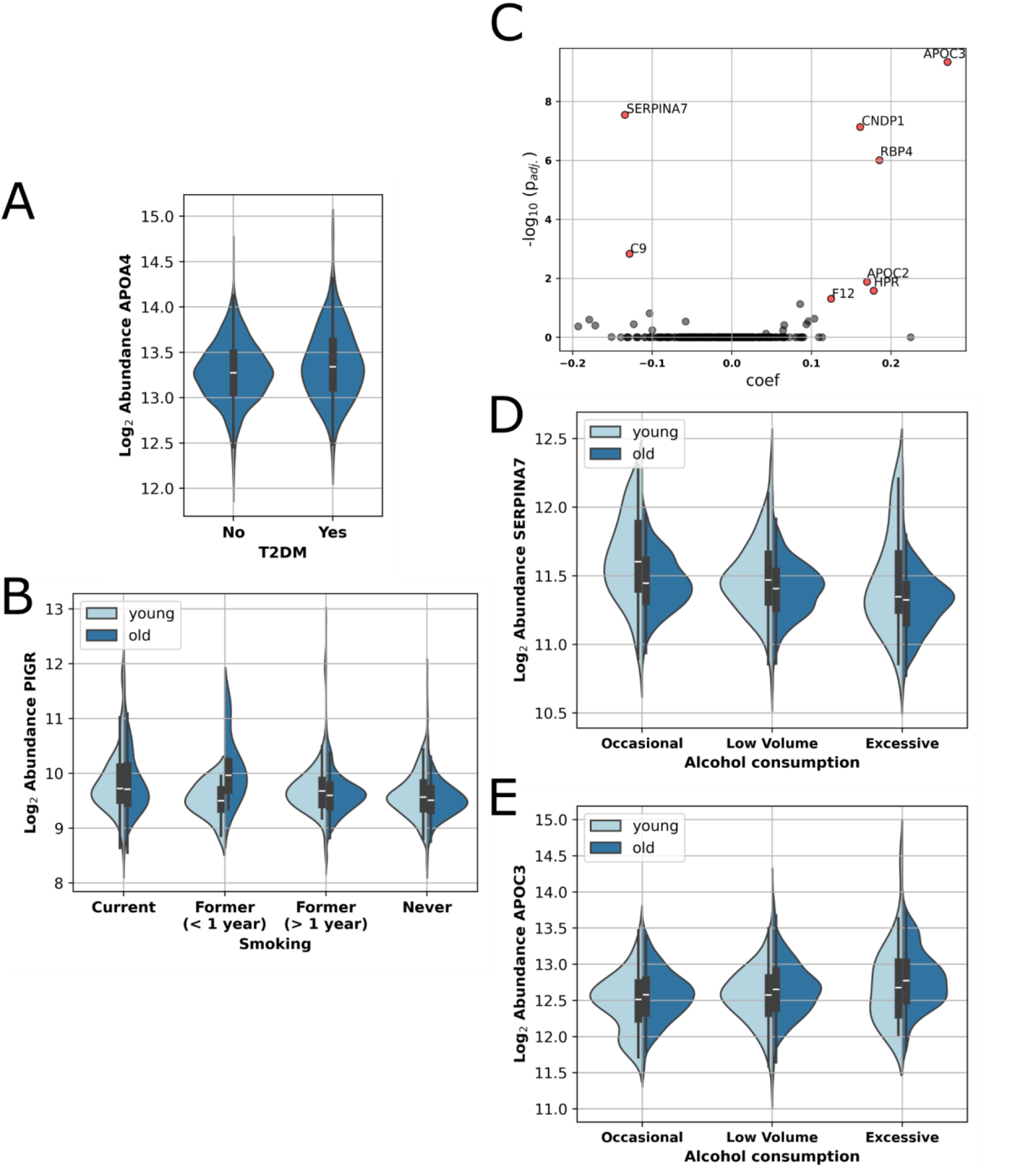
(A) Increased APOA4 abundance with age and with diabetes mellitus type II (T2DM) (B) Effect of smoking on the PIGR abundance levels. Coefficients and adjusted log10 p-values for the proteomic profiles across different participant groups (Volcano Plot) (C). Proteins significantly associated with alcohol consumption are marked in red and their gene names are annotated. Accompanying split violin plots depict decreasing SERPINA7 (D) and increasing APOC3 (E) levels in BASE-II participants with increasing alcohol consumption. Boxes in violin plots denote the 25% and 75% quantiles, and the median is displayed by a white line.

Next, we continued to address protein associations with metS in the entire cohort and type 2 diabetes in the older group. The abundance levels of four different apolipoproteins were found to be significantly associated with metS. Specifically, apolipoprotein D (APOD) and apolipoprotein A1 (APOA1) were decreased in participants diagnosed with metS, whereas apolipoprotein C2 (APOC2), and apolipoprotein C4 (APOC4) were increased (see Table S8).

Visceral fat accumulation is a key feature of metabolic syndrome and T2DM ^53,54^. We analyzed BMI-associated proteins in “overweight” and “obese” categories, considering T2DM and/or metabolic syndrome (metS) as covariates. Without lifestyle factors, 19 proteins were linked to “overweight” and/or “obese”BMI categories (see Table S3). Including T2DM as a covariate, dopamine-beta hydroxylase (DBH) was significantly associated with “obese”. In regression analyses with T2DM or metS, PGLYRP2 ^56^, SERPINA10 ^57^, and LYZ ^58^ were significant in both elevated BMI categories, which aligns with current literature. Fibulin 1 (FBLN1) remained significantly associated (p < 0.05) with both BMI categories, irrespective of covariates. The link between BMI and angiogenesis inhibitor FBLN1 downregulation has not been previously observed. Among the older participants N = 177, or 9.58% of all participants, were diagnosed with T2DM. This prevalence aligns closely with the previously reported rate of 9.9% for T2DM in Germany ^62^, based on statutory insurance data ^63^. Multiple linear regression analysis revealed a positive, significant association of APOA4 with T2DM. In an analysis of 539 participants diagnosed with metS, including some T2DM diagnosed participants, APOA4 (see Figure 3A) again emerged as the only significant protein whose variance exceeded technical variability when assessing biomarkers for T2DM.

Considering the established association between metS and T2DM, we performed multiple regression analysis on the complete cohort by including for metS and T2DM as covariates ^64,65^. The only protein that could exceed the technical threshold when both diseases are considered as covariates was APOB, which had a significant, positive association with metS but did not show a significant one for diabetes.

### Impact of menopausal hormone therapy on the high abundant serum proteome

We recently reported that HCU has a major impact on the high abundant fraction of the plasma proteome, in women under 40 years enrolled in the CHRIS study^21^, and as aforementioned, we detected a similar signature in the young women within the BASE-II cohort. With the focus on aging, we herein explored the effect of menopausal hormonal replacement therapy (MHT) in older women participating in the BASE-II. For this, we categorized the BASE-II women from the older group (N = 689, aged 61-84) into three groups: those who never underwent MHT (*never*, N = 295), those who completed MHT (*completed*, N = 359) and those who underwent treatment at sampling time (*ongoing*, N = 35). Only study participants with documented start and/or termination of MHT were considered. The median treatment duration for the MHT *completed* group was 11 years (IQR 8 years), and for the *ongoing* group, it was 18 years (IQR 9 years), reflecting a later onset of menopause ^70^. Subsequently, we employed multiple regression models incorporating age, BMI, and a categorical variable denoting MHT status to evaluate associations between protein abundances and the treatment.

We identified three proteins - angiotensinogen (AGT)^71–74^, peptidoglycan recognition protein 2 (PGLYRP2)^75^ and plasminogen (PLG) - to be significantly changed between study participants that are currently undergoing MHT treatment and those who have never received treatment. Additionally, we observed non-significant upregulation of ceruloplasmin (CP) and non-significant downregulation of vitamin K-dependent protein S (PROS1), C4b-binding protein alpha chain (C4BPA), lumican (LUM), and complement component 8A (C8A with MHT in our analysis.

Next, we investigated the predictive power of the serum proteome for classifying MHT status of the female BASE-II participants based on the serum proteome to enhance understanding of the underlying biological processes affected by hormonal treatments and add to already available literature concerning treatment effects on the serum proteome. We first matched female study participants under ongoing treatment (N = 35) to an equally sized control group by age and BMI based on propensity scores acquired from a generalized linear model ^76^ (Age and BMI demographics are listed in Table S7). Next, we selected plasminogen (PLG) and complement component 9 (C9) as the most informative features by a cross-validated, recursive feature elimination process^77^. We then trained a random forest ^78^ in a 5-Fold cross validation stratified by MHT status. The two-protein classifier achieved a mean test data area under the curve (AUC) of 0.82, comparable to the AUC of 0.80 for individual estradiol levels, the active ingredient in most MHT. Moreover, since a training AUC of 0.84 was obtained, the classifier should be robust against overfitting. ROC curves for different splits in the CV procedure can be found in Figure S258. Other hormones such as SHBG or testosterone were outperformed by the random forest predictor (Figure S259). To get a better understanding of the decision boundaries of the two proteins in the random forest classifier we predicted decision probabilities for the range of observed PLG and C9 fold changes (FC) to the log2 median abundance values (Figure 5B). The highest predictive probability for ongoing MHT was at PLG at FC above 0.2 and C9 log2 FCs below −0.1 or above 0.4, although the latter has to be interpreted with caution since it is not well displayed in our data. Conversely, we expect the highest predictive power for no MHT at PLG levels below 0.2 and C9 levels between −0.1 and 0.4. Ensemble models that rely on multiple weak learners rarely produce predicted probabilities close to 0 or 1 because it requires all the individual models to make the same decision. This is particularly unlikely in random forests due to the bootstrapping (random sampling with replacement) used to create each individual tree ^79^.

**Figure 4:**
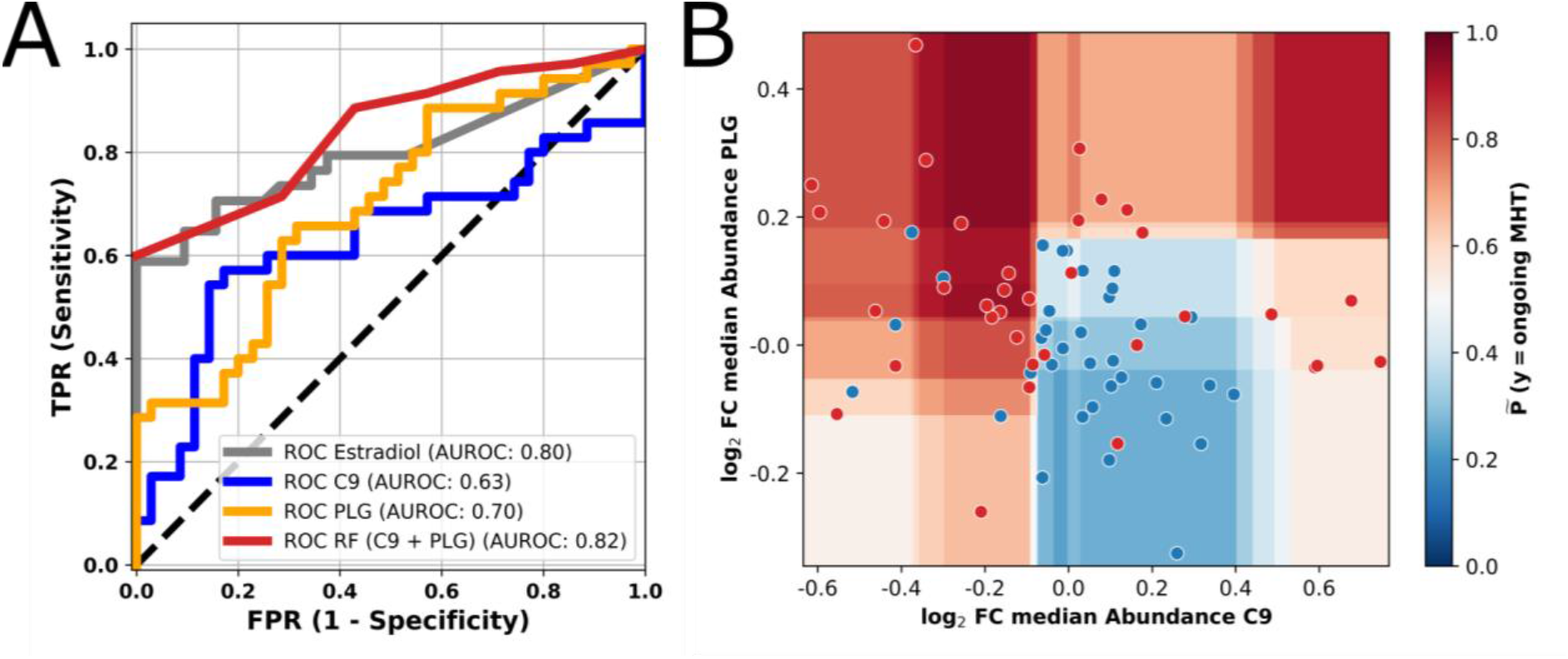
Machine learning classifier based on Complement Component 9 (C9) and Plasminogen (PLG) abundance levels slightly outperforms serum estradiol levels in predicting MHT status (A) Receiver operating characteristic (ROC) curve shows predictive performance of single parameter models with estradiol (coloured in gray), C9 (blue), PLG (orange) and the two protein random forest model (C9 + PLG) (red) predicting current hormonal treatment. (B) Contour plot depicting the predictive probability generated by the random forest classifier based on C9 and PLG levels. The areas highlighted in red represent the abundance ranges of PLG and C9, which support the classification of participants undergoing MHT. The added dots display the protein abundance measurements with participants currently under hormonal treatment (red) and controls (blue). Vertical and horizontal color gradations show cut-offs of the individual decision trees in the random forest model. Axis show the log2 FC to the median abundance of PLG (y) and C9 (x).

## Discussion

Large-scale serum and plasma proteome studies ^21,43,80,81^ have revealed disease prevalence, risk factors, and healthcare needs across diverse populations. These studies have identified putative biomarkers of aging ^1,21–24,80^, common lifestyle factors like Type II Diabetes, and administered drugs ^43,82,83^. Advancing age significantly heightens the risk for various chronic ailments alongside physical and cognitive decline, ultimately leading to mortality. Consequently, age directly impacts critical physiological processes, including inflammation ^84^, often referred to as ‘inflammaging’, and cellular senescence ^85^. However, biomarkers associated with aging have yet to gain widespread acceptance within the biomedical field, partly due to the non-trivial nature of validating them ^86^. The BASE-II ^14^ specifically aims to explore factors influencing a healthy aging process, presenting a rich resource for further investigations of age-related proteins.

In this study, mass spectrometry-based serum proteomics of 1859 BASE-II participants consistently quantified 2917 peptides from 248 protein groups at high precision. While the number of proteins in this upper fraction of the plasma proteome may seem limited, this group of proteins is secreted in many isoforms and is particularly interesting for biomedicine. Altogether, these proteins account for more than 99% of the plasma’s proteomic mass. This high abundant fraction covers many of the currently clinically used protein biomarkers, spans over many targets of FDA approved biomarkers, and closely reflects human physiology. Thereby they cover many functions in the complement system, coagulation, immunoglobulin, and acute-phase response ^10,15,87–91^.

Epidemiological covariates such as age, sex and BMI have a considerable impact on the general health of the population, but the mechanisms are multifactorial and complex. It has been shown that progressing age and very low or high BMI can pose a risk factor for multiple disease mechanisms and their progression ^92–94^. With the advances of personalized medicine, these factors are important for treatments tailored to individual patients ^95^. Specifically, the high abundant serum or plasma proteome that plays a vital role in disease pathology and metabolism, has to be investigated for these effects to reveal true relationships of treatment and disease ^9–12^. Furthermore, differences in epidemiological factors also pose a technical challenge when analyzing proteomic data. Common techniques to address this include balancing or using epidemiological information as covariates in linear (factorial) models ^96^.

Previous proteomic studies into aging cohorts successfully predicted age, mortality, and made attempts to distinguish biological from chronological age (‘proteomic clock’) ^5,22–24,97^. The BASE-II design is slightly different to these investigations, as sampling was not continuous over the age range, but rather categorical, and sampled from an aged group and a young control. The BASE-II is well annotated with demographic and metadata, which facilitated to probe for association of age with several demographic and physiological factors. Our association analysis did both, confirm proteins that were previously reported as age-dependent in the literature, as well as identify new proteins. Indeed, the abundance levels of 106 of 248 consistently quantified proteins were changed with age. Of these, 39% (N = 41) were not reported as age-associated in previous datasets ^21–30^. Thirty-two of these are immunoglobulin chains, which is consistent that the adaptive immune system changes as we age ^98^. We hope our results stimulate future investigations using antigen-capture technologies, to identify the biological targets of these immunoglobulins.

Furthermore, we report associations to some pathologically highly relevant proteins including the pro-inflammatory serum amyloid A4 (SAA4), which has been linked to various infectious diseases such as malaria or COVID19 ^99,100^ as well as rheumatic inflammation ^101^. Another protein identified as age-dependent in our study is proteoglycan 4 (PRG4), which has been associated with tissue regeneration ^31^ and inflammation regulation ^32,33^. It also functions as a boundary lubricant at the cartilage surface of joints, which function is known to decrease with progressing age. However, a direct connection to osteoarthritis, which can be caused due to decreased cartilage lubricants ^102^ are not yet described. In contrast to our findings, a negative regulation of PF4V1 and PPBP with age platelet activity has been described while platelet counts were shown to increase with age ^103^. Platelet factor 4 variant 1 (PF4V1)^104^ has been associated with angiogenesis, which, if pathogenic, can serve as a marker for autoimmune diseases, cancer, and cardiovascular diseases. Additionally, pro-platelet basic protein (PPBP) has been reported to have a wide range of functions, including neutrophil activation and antibacterial activity ^105^.

The sex dependency of the BASE-II proteome aligns well with the results obtained in other cohorts ^21,23,34–42^. The list of n = 23 sex dependent proteins includes the pregnancy zone protein (PZP), the sex hormone binding globulin (SHBG) and angiotensinogen (AGT). Moreover, as we have reported recently, HCU had the strongest impact on the serum proteome of BASE-II participants, with PGLYRP2 being the main suppressed protein, and AGT, SERPINA6, FETUB the most induced. We found new HCU associations for 15 proteins, such as the acute-phase protein SERPINA3 ^106^, the monocyte and macrophage cell surface protein CD14 ^107^ and the coagulation Factor 5 (F5).

Furthermore, we compared effect sizes for age, sex, BMI, and hormonal contraceptive usage with those from our recently presented analysis of the CHRIS dataset ^21^. We found correlations of up to 0.9 for HCU (Spearman’s rho), indicating a consistent effect on the plasma and serum proteome across both studies. Found age-associated proteins (N=53) in both studies included various components of the complement system (N=8) and apolipoproteins (5). Hence, the previously described age association of these mechanisms aligns with our results ^36,109,111–113^. The most prominent sex-dependent proteins in both the CHRIS and BASE-II proteomic datasets include SHBG, which is recognized as a key proteomic marker for sex differences, complement component 6 (C6), TTR, and ceruloplasmin (CP). Effect sizes for underweight BMI showed a negative correlation (−0.25), likely due to the small number of participants in this category (N=35), resulting in less consistent results.

We also addressed the role of body weight which is linked to various comorbidities ^114^. We did not find significant associations with underweight, which was most likely caused by the low sample size and hence lower statistical power. Similar to previous studies^115^, we detected an association of apolipoproteins D and M levels with elevated BMI. We also report novel protein associations with BMI including Apolipoprotein C2 (APOC2) and fibulin 1 (FBLN1), when corrected for metabolic disorders such as T2DM and metS. When both metabolic conditions (metS and T2DM) are considered as covariates, C5 emerges as a significantly upregulated protein in obese patients. This could be due to its role in regulating inflammation in adipose tissue ^116^, but also aligns well with our report, that the complement system is BMI dependent ^117^. Interestingly, this association is sensitive to metS and T2DM, which suggests that the onset of metabolic disease influences the complement system. Similarly, FN1 ^118^, VWF ^119^, ECM1 ^120^, and ADAMTS13 ^121^ are only significantly downregulated in obese and overweight patients when metS and T2DM are included as covariates. Furthermore, we found DBH significantly associated with “obese” BMI, when adjusted for T2DM in the older group of participants, aligning with prior findings in DBH-deficient mice ^55^.

BASE-II offers a valuable insight into proteomic signatures of metabolic disease like metS and T2DM as the incidence within our cohort aligns with previously reported national numbers ^63,122,123^. Our findings indicate that lifestyle factors like T2DM and metS diagnosis, influence the serum proteome to varying extents across different age groups. Notably, apolipoprotein A4 (APOA4) was significantly upregulated in T2DM cases, particularly when metS was considered. APOA4, primarily synthesized in the small intestine, is crucial for lipid metabolism, insulin sensitivity, and food intake regulation. Elevated APOA4 levels are also markers for kidney disease progression and diabetic nephropathy, ^124^ highlighting its potential as a marker for T2DM monitoring and prognosis. Significant associations were also found between metS and various apolipoproteins, including C2, C4, B, and D. Apolipoprotein D (APOD) was consistently downregulated, especially in the older cohort. APOD, involved in lipid trafficking and neuroprotection, enhances insulin sensitivity and metabolic health^125^ but is often reduced in metS patients, correlating with hyperchylomicronemia^126^, characterized by increased triglyceride and chylomicron levels in blood and dysfunctional APOD in metS ^127^. Notably, this downregulation was more pronounced among older metS patients, potentially reflecting an age-related increase in APOD expression ^128^. APOC2 ^52^ and APOC4, part of a gene cluster including APOE ^129^, are implicated in metabolic regulation. While APOE, a known marker of metabolic syndrome, was not significantly associated with metS in our study, APOC4 was upregulated in both age groups. APOC2 and APOC4 play pivotal roles in lipid metabolism and triglyceride processing associated with very low-density lipoprotein (VLDL) ^129,130^. Although the literature does not establish a direct link between APOC4 and metS, its role in lipid metabolism supports a potential relationship with metS and other cardiometabolic diseases. Prior research has also associated an increase in serum APOC2 levels with a diagnosis of metS ^52^. APOB serves as a clinical marker of atherosclerosis due to its association with low-density lipoprotein (LDL) and is an independent predictor of metS development ^127,131^. Previous research has shown that the clinical value of APOB in predicting metS is maintained even after accounting for the comorbidities linked with it ^127,132^. Our results further corroborate these findings, with significant upregulation of APOB in both age groups. Recent studies have highlighted the critical role of the apolipoprotein protein family in regulating triglycerides (TGs) and cholesterol metabolism, transport, and cellular management ^133^. These findings are consistent with our observations of deregulated apolipoprotein family members in metabolic diseases, underscoring their significance as potential therapeutic and diagnostic targets. APOB was the sole protein for which the abundance change associated with disease surpassed the technical threshold in the regression analysis that considered both T2DM and metS. Serum APOB levels are directly linked to insulin secretion ^134^, where decreased insulin sensitivity is associated with increased secretion and decreased clearance of APOB. Prediabetes, characterized by visceral fat accumulation and impaired insulin sensitivity, is considered an underlying condition of metS ^135^. The strong link between metS and T2DM has been established in relation to both incidence and the molecular mechanisms underlying their prognosis ^64,65^. Notably, administering oral antidiabetic drugs such as dipeptidyl peptidase-4 inhibitors (DPP4i) in T2DM patients was associated with reduced plasma APOB levels ^66^. Among the participants diagnosed with T2DM, 85 were receiving orally administered blood glucose-lowering drugs excluding insulin, which indicates the potential utility of APOB in monitoring treatment outcomes. Our study revealed a novel link between serum fibulin 1 (FBLN1) levels and higher BMI categories. FBLN1, a regulatory component of the extracellular matrix, is downregulated in higher BMI categories, indicating potential disruptions in cellular homeostasis, mobility, and adhesion ^138,139^. Markers of chronic inflammation and cellular degeneration such as lysozyme (LYZ) and alpha-2-macroglobulin (A2M) were also associated with individuals belonging to the “overweight” and “obese” categories, but these associations disappeared when adjusting for T2DM and metS. Only FBLN1 remained significantly linked to BMI, indicating its potential as a robust biomarker, regardless of T2DM or metS.

Lifestyle factors are modifiers of the blood protein profile. Plasma/serum proteomics might thus be able to complement patient questionnaires in clinical studies, as these have the reputation to be unreliable, specifically, when they address questions related to stigmatized lifestyle components, such as alcohol consumption or smoking ^140^. One notable marker for current and former smoking habits identified in our analysis was an increased abundance of PIGR in smokers. PIGR transports IgA through mucosal epithelia,^140^ and has been linked to smoking previously ^141,142^. In contrast to previous studies, we also report higher PIGR levels in some former smokers, specifically older participants who quit recently (less than a year) ^143^. Although imprecise responses about smoking habits in the questionnaires might partly explain these results (e.g., some individuals who identified as ex-smokers might still be smoking), the findings could also suggest that some individuals experience a longer recovery time from smoking-related chronic inflammation. Similarly, downregulation of IGGs, specifically IGHG2, relates to smoke-related inflammation ^143,144^. Our observations reinforce the idea of pulmonary inflammation as a consequence of smoking, which we show, might be traceable even in former smokers to a certain degree.

Excessive alcohol consumption (defined as more than 30 g/day) led to deregulated serum levels of several key metabolic proteins. The two with the highest significance, APOC3 and SERPINA7, were respectively upregulated and downregulated. Increased APOC3 levels with increased alcohol consumption have been previously observed ^145,146^ and decreased serum thyroxine as well as SERPINA7 levels have been linked to alcoholism ^147^. APOC3, a major HDL component released from hepatocytes, influences triglyceride catabolism and is associated with ethanol consumption, decreasing after alcohol withdrawal ^148,149^. Alcohol-induced increases in HDL are attributed to the increased expression of cholesteryl ester transfer protein in response to ethanol intake^150^. On the other hand, SERPINA7 was downregulated in excessive drinkers, a phenomenon observed in the context of the gut-thyroid axis ^151,152^. This observation could be attributed to liver dysfunction due to overworking, which impairs the liver’s ability to effectively synthesize SERPINA7^153^.

Eventually, we investigated the influence of MHT on the data subset of women from the older age group in the BASE-II. Statistical significant associations with MHT were found for peptidoglycan recognition protein 2 (PGLYRP2), angiotensinogen (AGT) and plasminogen (PLG), which have been described before to be influenced by estradiol-based hormone therapy or medication, such as hormonal contraceptives ^21,71,154,155^. Eight other proteins ^74,75,156,157^ showed trends similar to those described in the literature but did not reach significance levels. However, while other studies reported the dysregulation of Lp(A), APOA and APOB ^158,159^, these proteins were not identified in our analysis (Figure S257 and Table S6). Furthermore, no protein was found significant between women with previous MHT treatment and those that never received treatment. Thus, in our dataset, we did not find any significant long lasting changes in the plasma proteome by MHT. The impact of MHT on the proteome (with N = 3 significant protein associations) compared to HCU (with N = 55) aligns well with the lower dosage of estradiol in MHT and the predominant use of bioidentical estradiol as an active ingredient in MHT compared to classical HCU, which predominantly contains the more powerful synthetic ethinylestradiol and much higher dosages. Employing machine learning, we successfully differentiated between participants who had undergone MHT during sampling from those who had not, achieving an AUROC of 0.82%. Interestingly, we obtained best separation with a random forest model trained solely on abundance levels of PLG and C9, which were identified beforehand in a recursive feature elimination process, even slightly outperforming the predictive performance of estradiol (AUROC: 0.8), a major component of most used MHTs.

## Conclusion

In summary, our study reports a significant serum proteome remodeling during aging. Our data confirms previous, and reports new associations, of serum proteins that change during aging dependent on age, sex, and BMI, and highlights the influence of lifestyle factors such as smoking and alcohol consumption. In particular, our findings underscore the impact of hormonal therapies, HCU in young female study participants, and MHT in older female participants, which both have a quantitative impact on the serum proteome. Especially HCU had a stronger impact than any other covariates on the serum proteome in the younger female participants. Our study did not assess whether the proteomic changes resulting from HCU or MHT are related to health outcomes. We hope however our study will stimulate studies to clarify this question. Overall, our findings provide valuable insights into the factors influencing serum proteome remodeling during aging and pave the way for future research to explore the health implications and potential therapeutic interventions.

## Limitations

The study design includes two distinct age groups (27-37 and 60-84 years) and therefore we cannot claim to ultimately depict effects between these age ranges. However the high overlap of age-associated proteins between our analysis and comparable studies ^21–24^ implies that our analysis is reliable. The analysis of MHT effects is based on a relatively small sample size of positive cases (N = 35). To minimize adverse effects as much as possible, we therefore matched the treatment group to a control group with comparable age and BMI. The small sample size also limits an analysis of admission routes and active drug ingredients (Gestagen and Estrogen) of MHT. But we report this information, for future reference in the supplement (see Figure S260). Another potential limitation of our study is the categorization of alcohol consumption based on self-reported daily intake. Although we used the same thresholds for both men and women as cited in previous studies ^160–162^ (see Methods), the German Center for Addiction Issues (DHS) defines low-risk alcohol consumption differently for women (<12 g/day) and men (<24 g/day)^163^. These differing limits complicated the definition of a moderate alcohol consumption group and led to a larger portion of the cohort being classified as excessive drinkers. Given the distribution of age and sex among participants as well as known clinical details about the group, which were also covariates in our analysis, we chose to define alcohol consumption limits based on established studies.

## Methods

### Berlin Aging Study II

Berlin Aging Study II (BASE-II) participants were recruited from the greater metropolitan area of Berlin, Germany. The multidisciplinary BASE-II aims to identify mechanisms and factors contributing to healthy vs unhealthy aging. In total 1,671 older participants (aged ≥60 years) and 500 younger (20-37 years) were assessed in the medical part of the study at baseline between 2009 and 2014 ^14^. The medical assessments were conducted in accordance with the Declaration of Helsinki and approved by the Ethics Committee of the Charité – Universitätsmedizin Berlin (approval number EA2/029/09) and were registered in the German Clinical Trials Registry as DRKS00009277. Blood was drawn from all participants after an >8h fasting period and kept at 4–8 °C until analysis on the same day or was stored at −80 °C (serum, plasma). Estradiol, total testosterone, and sex hormone-binding globulin (SHBG) were measured in serum probes by fluoro-, radio- and electrochemiluminescence immunoassays by an accredited laboratory. Standard laboratory parameters (including estradiol, testosterone and SHBG) and proteome analyses were performed on blood from the same draw. Smoking behavior (current smoker, former smoker, never smoker), hormonal contraceptive usage and menopause hormone replacement therapy were assessed during the 1:1 interview by trained study personnel. Alcohol consumption was assessed in g/day via food frequency questionnaire ^164^. Body Mass Index (BMI, kg/m2) was calculated from height and weight measured with an electronic measuring station (seca 763 measuring station, SECA, Germany). Diabetes mellitus type 2 was diagnosed based on American Diabetes Association (ADA) guidelines ^165^, and the metabolic syndrome was diagnosed according to Alberti et al. ^166^.

### Sample Preparation

In this study, a total of 2,191 samples were subjected to measurement. Among these, 105 standardized, commercially available serum set samples (SER-SPL, Zen-Bio, Durham, NC, USA) and 96 were pooled study samples that were utilized to monitor measurement quality and control technical variation. Measurements were performed in 8 batches, each consisting of 3 96-well plates. Serum samples were randomly distributed across all plates.

Semi-automated in-solution digestion was performed as previously described for high throughput clinical proteomics^15^. All stocks and stock plates were prepared in advance to reduce variability and were stored at −80°C until use. Briefly, 5 μl of thawed samples were transferred to the denaturation and reduction solution (55 μl 8 M Urea, 100 mM ammonium bicarbonate (ABC), 5 mM dithiothreitol per well) mixed and incubated at 30°C for 60 minutes. Five microliters were then transferred from the iodoacetamide stock solution plate (100 mM) to the sample plate and incubated in the dark at RT for 30 minutes before dilution with 100 mM ABC buffer (340 μl). 220 μl of this solution was transferred to the pre-made trypsin stock solution plate (12.5 μl, 0.1 μg/μl) and incubated at 37°C for 17 h (Benchmark Scientific Incu-Mixer MP4). The digestion was quenched by addition of formic acid (10% v/v, 25 μl) and cleaned using C18 solid phase extraction in 96-well plates (BioPureSPE Macro 96-Well, 100 mg PROTO C18, The Nest Group). The eluent was dried under vacuum and reconstituted in 60 μl 0.1% formic acid. Insoluble particles were removed by centrifugation and the samples transferred to a new plate.

### Liquid Chromatography and Mass Spectrometry

Digested Peptides were analyzed on a Bruker timsTOF Pro mass spectrometer coupled to a Bruker VIP-HESI electrospray source. Chromatographic separation was performed on the same column type but with only 2 µg and a flow rate of 500 µL/min. For diaPASEF acquisition ^167^, the electrospray source was operated at 3000 V of capillary voltage, a drying gas flow rate of 10 L/min and 240 °C. The diaPASEF windows scheme was set up as followed: we sampled an The ion mobility range was sampled from 1/K0 = 1.30 to 0.7 Vs/cm2 using ion accumulation times of 100ms and ramp times of 133ms in the dual TIMS analyzer, with each cycle times of 1.25 s. The collision energy was lowered as a function of increasing ion mobility from 59 eV at 1/K0 = 1.6 Vs/cm2 to 20 eV at 1/K0 = 0.6 Vs/cm²2. For all experiments, TIMS elution voltages were calibrated linearly to obtain the reduced ion mobility coefficients (1/K0) using three Agilent ESI-L Tuning Mix ions (m/z, 1/K0: 622.0289, 0.9848 Vs/cm2; 922.0097, 1.1895 Vs/cm2; and 1221.9906, 1.3820 Vs/cm2).

### Data processing

Raw MS data was processed using DIA-NN v1.8.1 ^19^. DIA-NN mass accuracies and scan window size were fixed to ensure reproducibility (scan window size: 7; MS1/MS2 15 ppm). An external, publicly available spectral library was used for all measurements^20^. The spectral library was annotated using the Human UniProt^168^ isoform sequence database (Proteome ID: 3AUP000005640)^20^. It is accessible via the PRIDE database under ID PXD013231.

All preprocessing steps (including normalization, batch correction and imputation) were performed in the R programming language (v4.3.1). The raw Dia-NN output was imported into R using the *msdap* package^169^ and initially filtered for serum set QC samples and non-proteotypic peptides (peptides mapping to multiple proteins).

Subsequently, the dataset underwent cyclic loess normalization^170^ to normalize peptide intensities between each sample and each other sample, followed by the ‘modebetween_protein’ method that performs the same normalization on a protein level. The normalized data was filtered iteratively to remove samples with too many missing peptide intensities (3 median absolute deviations), followed by the filtration of low-presence peptides measured in less than 30% of all samples. The filtered data was then imputed using Bayesian PCA (bpca) from R’s *pcaMethods* package^171^, where the missing peptide intensities were imputed within the same batch, sex (male/female) and age-group combinations (young/old), but only for peptides in those combinations which were measured in at least 66% of the samples. Subsequently, batch effects were corrected between all 24 plates using the *removeBatchEffect* function in the *limma* Bioconductor package^172^.

Finally, any remaining missing values in precursors with less than or equal to 34% missing values were imputed using the *impute.knn* k-nearest neighbor (knn) algorithm from package impute version 1.76 ^173^, followed by a protein-level summarisation using the function *rcModelPLM* probe-level model (plm) from package preprocessCore version 1.64^174^. The final dataset after preprocessing consists of 248 proteins and 2917 Peptides for 2079 samples. However, the number of proteins includes multiple isoforms for Kinogen 1 (2 isoforms, KNG1), Inter-alpha-trypsin inhibitor heavy chain H4 (2, ITIH4), Gelsolin (2, GSN), alpha-1-B glycoprotein (2, A1BG) and Fibulin 1 (3, FBLN1). Peptide Sequences for all proteins are reported in Tables S3-S5.

### Statistical Analysis and Machine Learning

Functional protein analysis was performed with the PANTHER classification system^175^. Subsequent statistical analysis and machine learning of protein data was conducted in the Python programming language (v3.11), unless specified otherwise. Principal Component Analysis was conducted with standardized protein abundance data by employing the scikit-learn package^176^. The arrows added to the plot indicate both the direction and magnitude of the loadings. A significance level of alpha <= 0.05 was employed and p-values were adjusted by bonferroni. Additionally, we employed a second significance criteria to account for technical variance, by comparing the coefficient of categorical covariates as a percentage to the CV of the study pools. We excluded 30 samples due to missing age or BMI information and one sample due to a duplicated measurement. Linear models, as implemented in statsmodels^177^ ordinary least squares (OLS) function, were fitted to the log2 protein abundance of all proteins to investigate associations. Age was treated as a continuous variable and divided by 10, thus the age-related coefficients show the change in 10 years difference. All other variables used in the association analysis were treated as categorical variables and were assigned a fixed reference category. For linear modeling, defined the BMI category “healthy” (18.5 < BMI ≤ 25; reference) as the baseline level and obtained coefficients for the contrast with “underweight” (BMI < 18.5), “overweight” (25 < BMI ≤ 30) and “obese” (BMI > 30). Smoking was also categorized in 4 groups, such as never smoked (never), stopped more than a year ago (former > 1 year), less than year ago (former < 1 year) and current smokers (current). The alcohol consumption of participants was categorized according to defined ranges in the related literature ^160–162^. In these studies, occasional drinking is defined as consuming less than 1.3 g of ethanol per day, low volume drinking is defined as consuming between 1.3 g and 30 g of ethanol per day, and excessive alcohol consumption is defined as consuming more than 30 g of ethanol per day. Furthermore, reference categories for sex, MHT, metS and diabetes were men, never treated with MHT and not diagnosed for metS and diabetes, respectively. Standardized effect sizes (ES) were obtained by subtracting the mean of every protein and dividing through its standard deviation prior to linear modeling. Spearman’s rho was used to calculate correlation coefficients as implemented in Scipy^178^ if not indicated otherwise. Hormonal contraceptive usage (HCU) was defined as in intake within the last 3 months and considered only oral and not topical administration. Protein CVs for study, pooled and standardized serum set samples can be found in Table S1. CVs were calculated by dividing the standard deviation of the abundance data by its mean and expressed as a percentage.

Prior to machine learning, we constructed an age and BMI matched control group (N=35) of women that were never treated with any kind of hormonal replacement therapy to ensure a balanced dataset. Matching was conducted by employing R’s matchit package^76^ with a “nearest” matching method and by estimating propensity scores by a “mahalanobis” based distance matrix. Demographics of both, control and treatment group, are shown in Table S7. We employed random forest models to classify women based on their MHT status as implemented in the scikit-learn^176^ package. Next, to the best predictive performance compared to other machine learning architectures such as Support Vector machines or boosting algorithms, random forests also do not need additional feature scaling and are robust to noise and overfitting due to bootstrapping. To reduce model complexity and only select the most informative features, we employed a cross-validated recursive feature elimination approach (as implemented in scikit-learn) with 10 features removed at every iteration. We finetuned the model using grid search, which constrained the model to have 10 trees and a maximum node depth of 2. Prediction scores were obtained in a 5-Fold cross validation procedure, where training and test sets were balanced by MHT status. To obtain predicted probabilities for Figure 5B, we trained a random forest with identical hyperparameters on all 70 individuals that have been used in the cross validation before. The predicted probabilities of random forest models are calculated by dividing the number of votes for a particular class by the total number of trees. Receiver Operating Curves (ROC), the area under the receiver operating curve (AUROC) were calculated with scikit-learn. To evaluate model performance across multiple cross validation sets, we aggregated the false positive rates (FPR) from each test set, creating a comprehensive set of unique and sorted FPR values. We then interpolated the true positive rates (TPR) for these FPR values from each test set, averaged the interpolated TPRs, and calculated the mean ROC curve.

## Data Availability

The raw mass spectrometry data of study pools and standardized, commercially available serum set samples, as well as the raw DIA-NN output and the FASTA file used for spectral library annotation, will be deposited in the ProteomeXchange Consortium via the PRIDE partner repository upon publication. To comply with data privacy rules, accessing human data requires a request to the access committee. Please contact the scientific coordinator, Ludmila Mueller, at for additional information about the procedures.

## Supplementary Material

PDF document contains Supplementary Figures S1-S260 and Supplementary Tables S1 and S6-S12. Spreadsheets (xlsx) with Supplementary Table S2 contains functional annotations of protein groups present in the dataset. Tables S3-S5 contain results of association analysis for the whole cohort and the younger and older age group, respectively.

## Data Availability Statement

BASE-II proteome data is available only upon reasonable request. Please contact Ludmila Müller, scientific coordinator, at, for additional information. The raw mass spectrometry data of study pools and standardized, commercially available serum set sample as well as the raw DIA-NN output and used FASTA file for spectral library annotation will be deposited to the ProteomeXchange Consortium via the PRIDE partner repository^179^ upon publication.

## Acknowledgements

The authors of this paper thank all study participants of the Berlin Aging Study II, as well as all people involved in planning this study, executing sample collection, conducting questionnaire survey and preparing the data. We thank the Charité Core Facility High Throughput Mass Spectrometry for conducting the measurements. We thank Vivien Bahr for clinical background information regarding glucose lowering drugs and Oliver Lemke for inspiration on how to visualize a 2-feature random forest.

## Ethics: Institutional Review Board Statement

All participants gave written informed consent. The Ethics Committee of the Charité – Universitätsmedizin Berlin approved the study (approval number EA2/029/09). The study was conducted in accordance with the Declaration of Helsinki and was registered in the German Clinical Trials Registry as DRKS00009277.

## Author Contributions

Conceptualization: M.R., I.D., C.D., R.S.M.; Investigation: K.T., D.L., O.S., V.F., M.M.; Formal Analysis: C.D., R.S.M., O.S., V.F., M.M., J.R.; Writing – original draft preparation: C.D., R.S.M., O.S., K.T.; Writing – review and editing: C.D., R.S.M., O.S., J.R., I.D., M.R.; Funding acquisition: I.D., M.R.

