## Supplementary material for "Menopause Hormone Replacement Therapy and Lifestyle Factors affect Metabolism and Immune System in the Serum Proteome of Aging Individuals": Supp_Information

June 21, 2024

#### Contents

**1 Technical Parameter and Dataset Overview** **2**
1.1 Protein abundance distributions . . . . . . . . . . . . . . . . . . . . . . . . . . . . 4

**2 Correlation between Protein Abundance and Diagnostic Assays** **67**

**3 Additional Excel Tables** **68**

**4 Effect Size Concurrence between CHRIS and BASE II** **70**

**5 Influence of MHT on the Serum Proteome of BASE II Participants** **71**

**6 Association Analysis for Lifestyle Factors and Comorbidities** **76**

### 1 Technical Parameter and Dataset Overview

**Table S1:** Number of participants, proteins and peptides as well as coefficient of variation (CV in %) for study samples, pooled samples and commercial serum samples at different stages of the preprocessing.

| **Stage** | $N_{participants}$ (%) | $N_{Proteins}$ (%) | $N_{Peptides}$ (%) | $CV_{study}$ | $CV_{pools}$ | $CV_{comm.}$ |
|---|---|---|---|---|---|---|
| I | 2192 (100) | 5640 (100) | 410 (100) | 56.9 | 27.6 | 35.1 |
| II | 2192 (100) | 5640 (100) | 410 (100) | 45.3 | 25.1 | 30.6 |
| III | 2089 (95.3) | 4386 (77.8) | 349 (85.1) | 42 | 23.3 | 29.6 |
| IV | 2089 (95.3) | 4386 (77.8) | 349 (85.1) | 50.3 | 26.61 | 34.2 |
| V | 2089 (95.3) | 4386 (77.8) | 349 (85.1) | 37.4 | 22.9 | 26.5 |
| VI | 2079 (94.8) | 2917 (51.7) | 248 (60.49) | 27.8 | 10.9 | 12.7 |

**Stages of preprocessing process**

I. **Initial data set:** Initial set of samples and proteins, excluding commercial samples (SP.QC plasma and serum) and non-proteotypic peptides (peptides mapping to multiple proteins)

II. **Initial normalization:** Data set after first normalization step

III. **Filtration:** Data set without outlier samples (many missing values) and low-presence peptides (peptides with less than 30% presence in all samples)

IV. **Imputation** based on bayesian PCA

V. **Batch correction** on peptide level

VI. **Imputation and normalization** KNN imputation and cyclic loess normalization

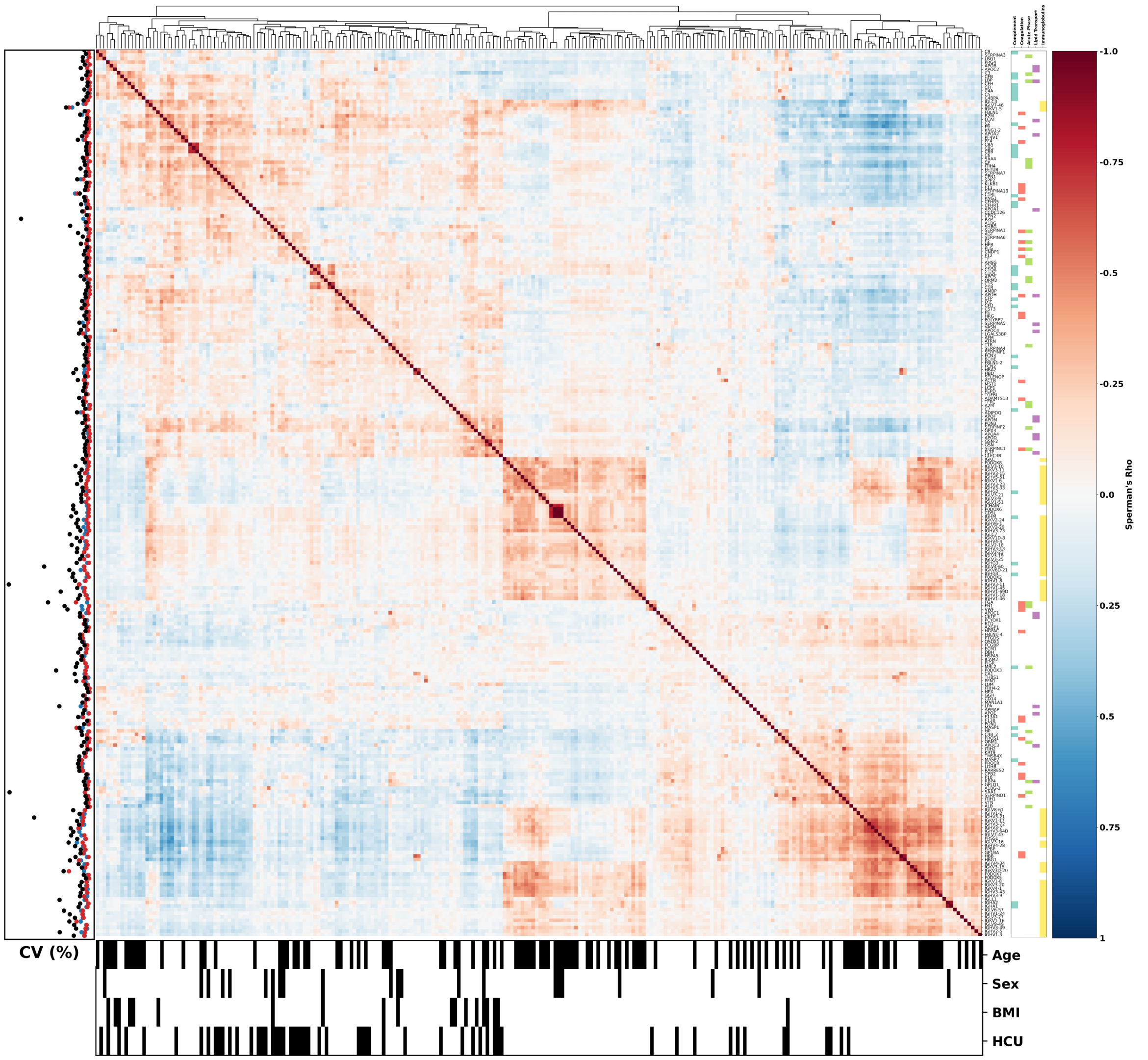


**Figure S1:** Hierarchically clustered heatmap depicts spearman's rho correlation between the 248 protein groups of the data set. The left panel depicts the coefficient of variation (CV) for study samples (black), pooled samples (red) and standardized serum samples (blue), showing overall higher CV for study samples. The lower panel depicts assoications with age, sex, BMI and hormonal contraceptive usage (HCU) in the analysis over the whole cohort. Right panel depicts annotations to several functional groups (Complement System, Coagulation, Acute-Phase, Lipid Transport, Immunoglobulins) based on gene ontology (GO) terms.

#### 1.1 Protein abundance distributions

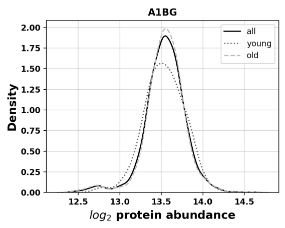


**Figure S2:** $\log_2$ Abundance distribution of A1BG

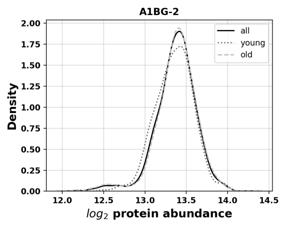


**Figure S3:** $\log_2$ Abundance distribution of A1BG-2

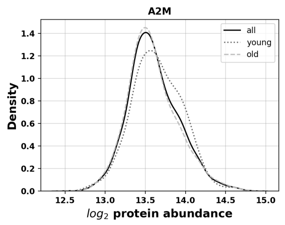


**Figure S4:** $\log_2$ Abundance distribution of A2M

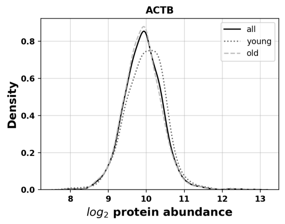


**Figure S5:** $\log_2$ Abundance distribution of ACTB

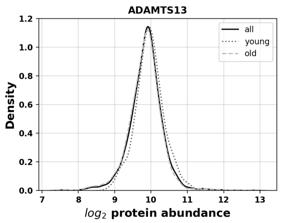


**Figure S6:** $\log_2$ Abundance distribution of ADAMTS13

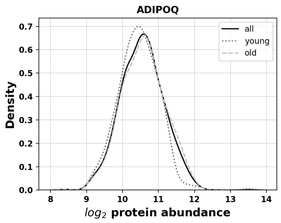


**Figure S7:** $\log_2$ Abundance distribution of ADIPOQ

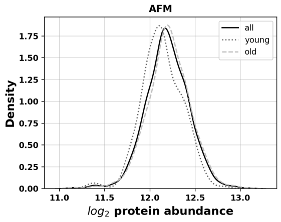


**Figure S8:** $\log_2$ Abundance distribution of AFM

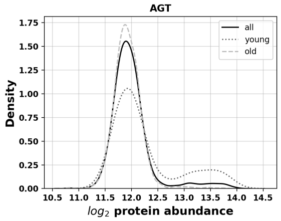


**Figure S9:** $\log_2$ Abundance distribution of AGT

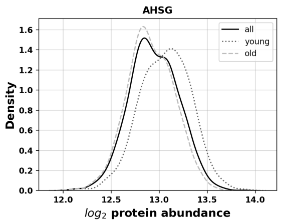


**Figure S10:** $\log_2$ Abundance distribution of AHSG

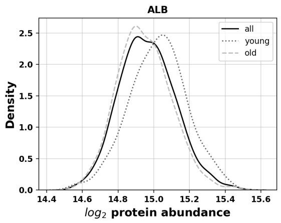


**Figure S11:** $\log_2$ Abundance distribution of ALB

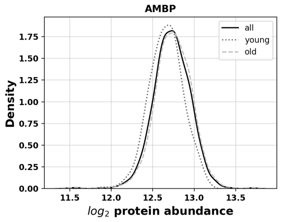


**Figure S12:** $\log_2$ Abundance distribution of AMBP

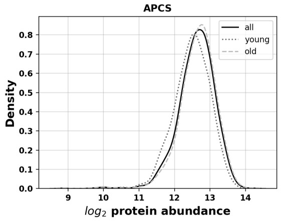


**Figure S13:** $\log_2$ Abundance distribution of APCS

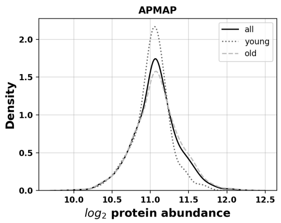


**Figure S14:** $\log_2$ Abundance distribution of APMAP

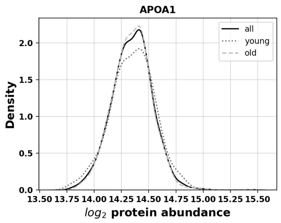


**Figure S15:** $\log_2$ Abundance distribution of APOA1

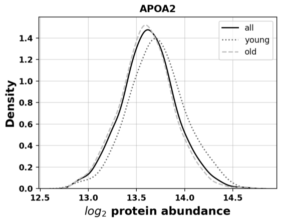


**Figure S16:** $\log_2$ Abundance distribution of APOA2

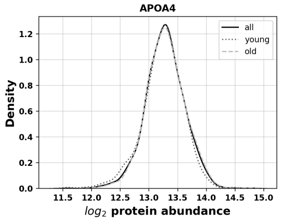


**Figure S17:** $\log_2$ Abundance distribution of APOA4

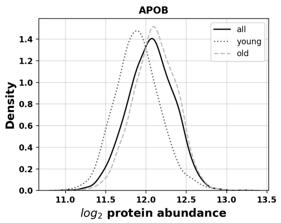


**Figure S18:** $\log_2$ Abundance distribution of APOB

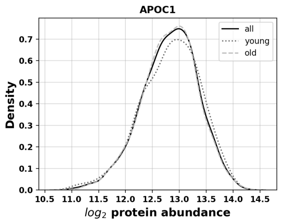


**Figure S19:** $\log_2$ Abundance distribution of APOC1

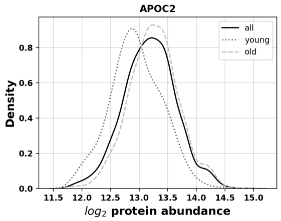


**Figure S20:** $\log_2$ Abundance distribution of APOC2

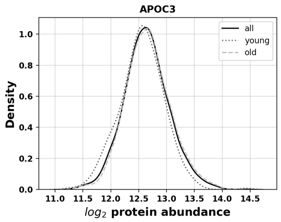


**Figure S21:** $\log_2$ Abundance distribution of APOC3

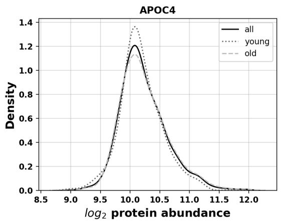


**Figure S22:** $\log_2$ Abundance distribution of APOC4

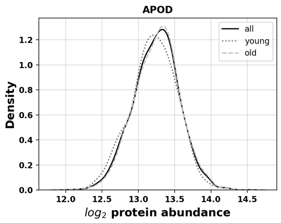


**Figure S23:** $\log_2$ Abundance distribution of APOD

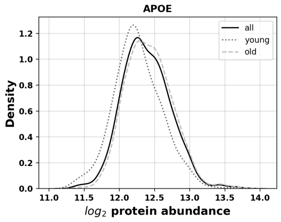


**Figure S24:** $\log_2$ Abundance distribution of APOE

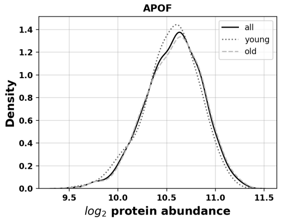


**Figure S25:** $\log_2$ Abundance distribution of APOF

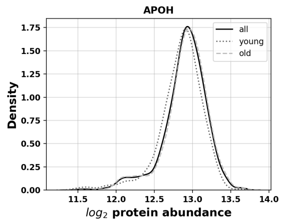


**Figure S26:** $\log_2$ Abundance distribution of APOH

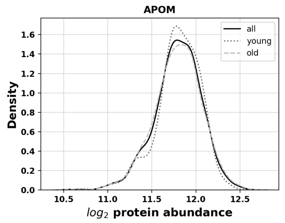


**Figure S27:** $\log_2$ Abundance distribution of APOM

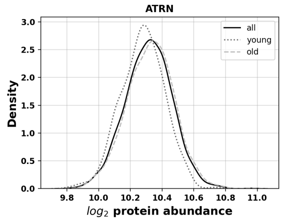


**Figure S28:** $\log_2$ Abundance distribution of ATRN

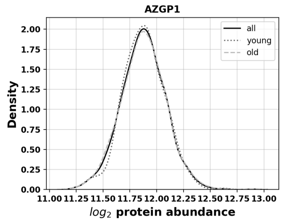


**Figure S29:** $\log_2$ Abundance distribution of AZGP1

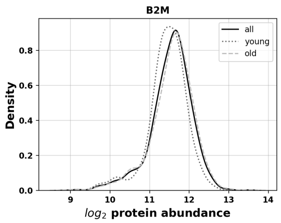


**Figure S30:** $\log_2$ Abundance distribution of B2M

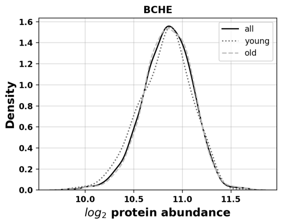


**Figure S31:** $\log_2$ Abundance distribution of BCHE

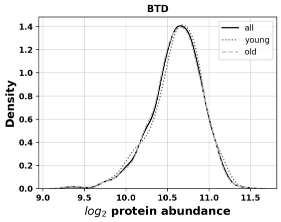


**Figure S32:** $\log_2$ Abundance distribution of BTD

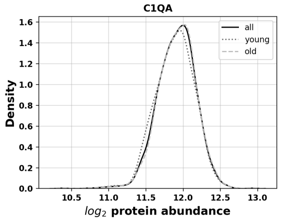


**Figure S33:** $\log_2$ Abundance distribution of C1QA

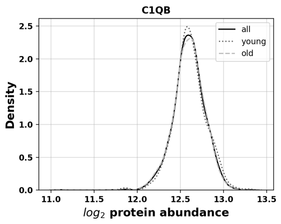


**Figure S34:** $\log_2$ Abundance distribution of C1QB

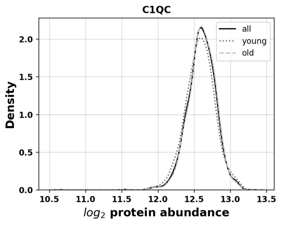


**Figure S35:** $\log_2$ Abundance distribution of C1QC

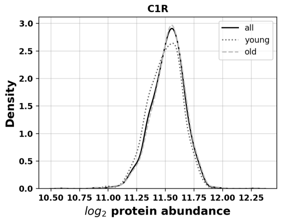


**Figure S36:** $\log_2$ Abundance distribution of C1R

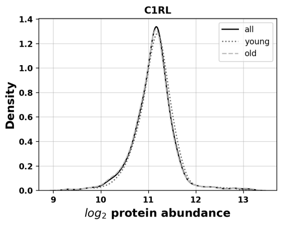


**Figure S37:** $\log_2$ Abundance distribution of C1RL

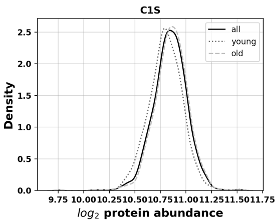


**Figure S38:** $\log_2$ Abundance distribution of C1S

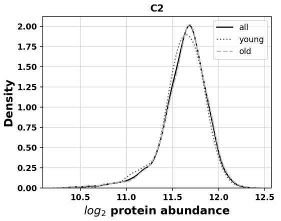


**Figure S39:** $\log_2$ Abundance distribution of C2

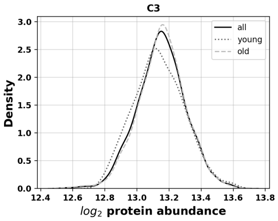


**Figure S40:** $\log_2$ Abundance distribution of C3

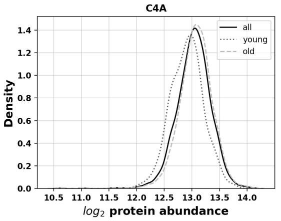


**Figure S41:** $\log_2$ Abundance distribution of C4A

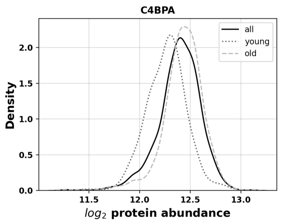


**Figure S42:** $\log_2$ Abundance distribution of C4BPA

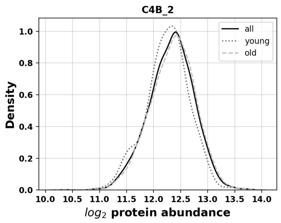


**Figure S43:** $\log_2$ Abundance distribution of $C4B_2$

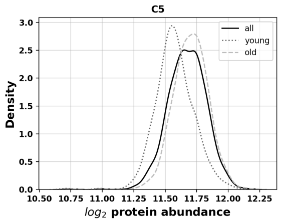


**Figure S44:** $\log_2$ Abundance distribution of C5

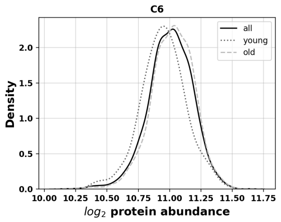


**Figure S45:** $\log_2$ Abundance distribution of C6

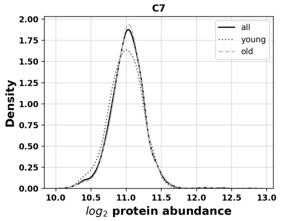


**Figure S46:** $\log_2$ Abundance distribution of C7

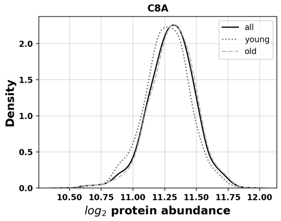


**Figure S47:** $\log_2$ Abundance distribution of C8A

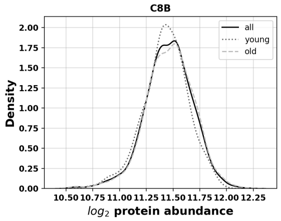


**Figure S48:** $\log_2$ Abundance distribution of C8B

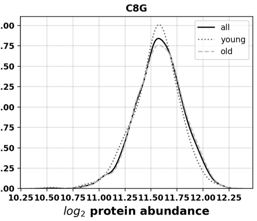


**Figure S49:** $\log_2$ Abundance distribution of C8G

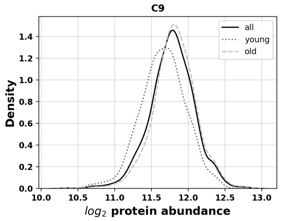


**Figure S50:** $\log_2$ Abundance distribution of C9

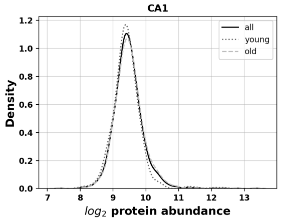


**Figure S51:** $\log_2$ Abundance distribution of CA1

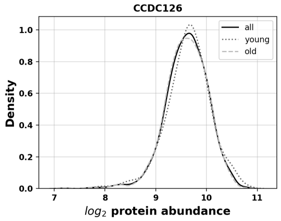


**Figure S52:** $\log_2$ Abundance distribution of CCDC126

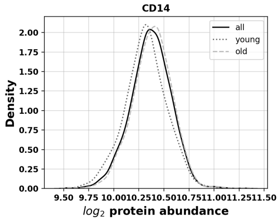


**Figure S53:** $\log_2$ Abundance distribution of CD14

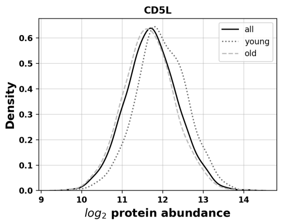


**Figure S54:** $\log_2$ Abundance distribution of CD5L

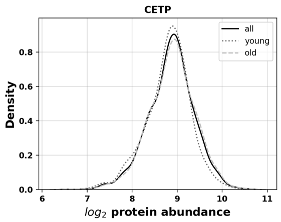


**Figure S55:** $\log_2$ Abundance distribution of CETP

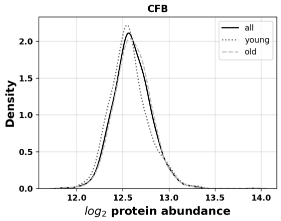


**Figure S56:** $\log_2$ Abundance distribution of CFB

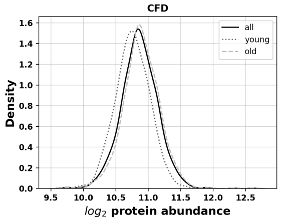


**Figure S57:** $\log_2$ Abundance distribution of CFD

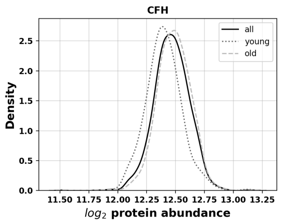


**Figure S58:** $\log_2$ Abundance distribution of CFH

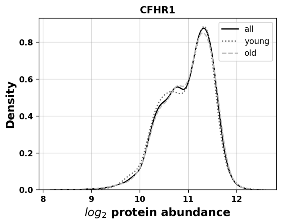


**Figure S59:** $\log_2$ Abundance distribution of CFHR1

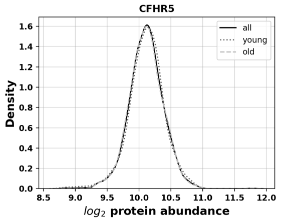


**Figure S60:** $\log_2$ Abundance distribution of CFHR5

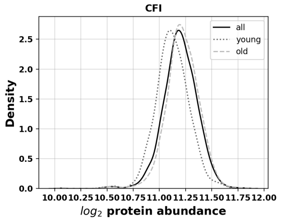


**Figure S61:** $\log_2$ Abundance distribution of CFI

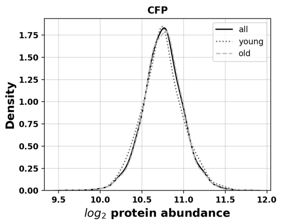


**Figure S62:** $\log_2$ Abundance distribution of CFP

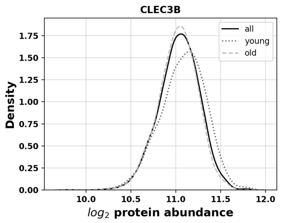


**Figure S63:** $\log_2$ Abundance distribution of CLEC3B

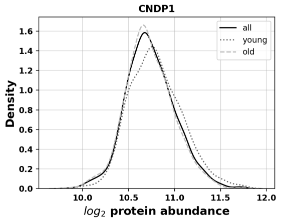


**Figure S64:** $\log_2$ Abundance distribution of CNDP1

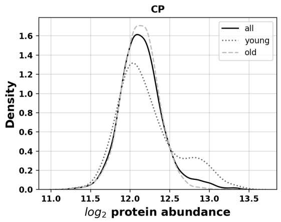


**Figure S65:** $\log_2$ Abundance distribution of CP

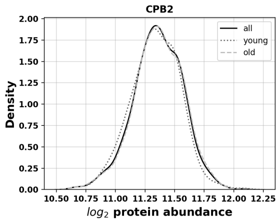


**Figure S66:** $\log_2$ Abundance distribution of CPB2

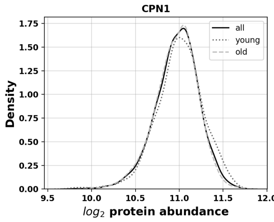


**Figure S67:** $\log_2$ Abundance distribution of CPN1

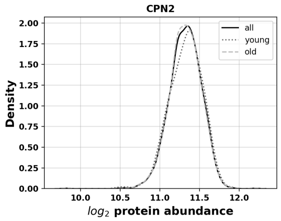


**Figure S68:** $\log_2$ Abundance distribution of CPN2

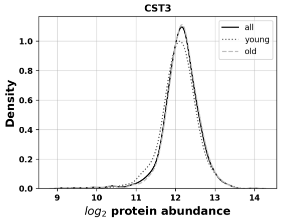


**Figure S69:** $\log_2$ Abundance distribution of CST3

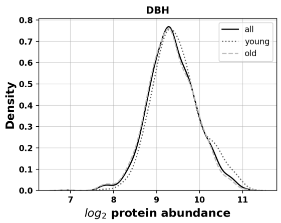


**Figure S70:** $\log_2$ Abundance distribution of DBH

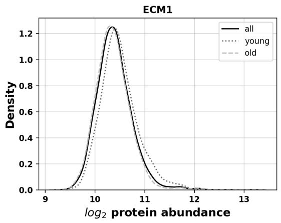


**Figure S71:** $\log_2$ Abundance distribution of ECM1

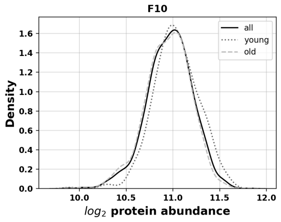


**Figure S72:** $\log_2$ Abundance distribution of F10

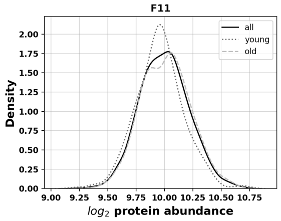


**Figure S73:** $\log_2$ Abundance distribution of F11

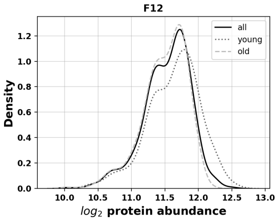


**Figure S74:** $\log_2$ Abundance distribution of F12

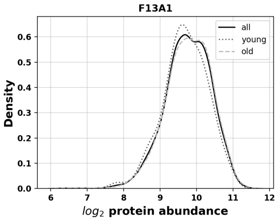


**Figure S75:** $\log_2$ Abundance distribution of F13A1

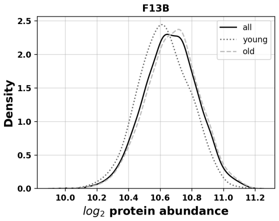


**Figure S76:** $\log_2$ Abundance distribution of F13B

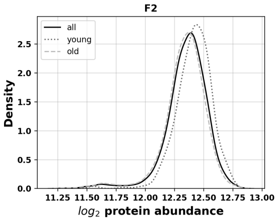


**Figure S77:** $\log_2$ Abundance distribution of F2

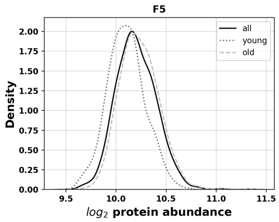


**Figure S78:** $\log_2$ Abundance distribution of F5

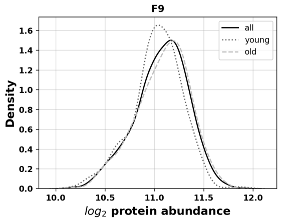


**Figure S79:** $\log_2$ Abundance distribution of F9

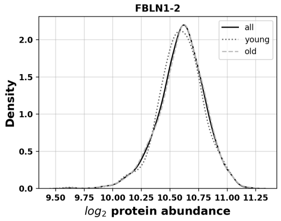


**Figure S80:** $\log_2$ Abundance distribution of FBLN1-2

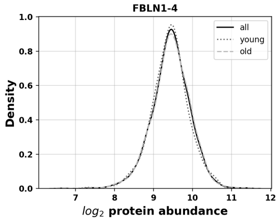


**Figure S81:** $\log_2$ Abundance distribution of FBLN1-4

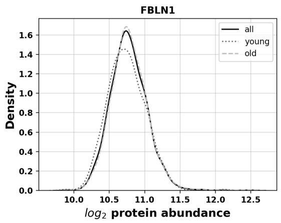


**Figure S82:** $\log_2$ Abundance distribution of FBLN1

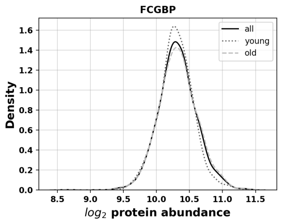


**Figure S83:** $\log_2$ Abundance distribution of FCGBP

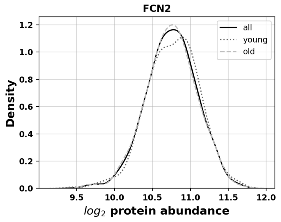


**Figure S84:** $\log_2$ Abundance distribution of FCN2

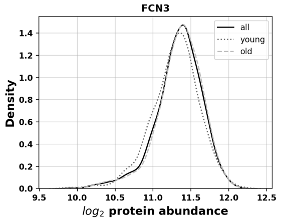


**Figure S85:** $\log_2$ Abundance distribution of FCN3

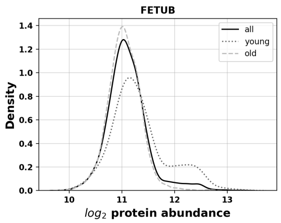


**Figure S86:** $\log_2$ Abundance distribution of FETUB

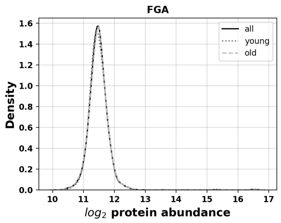


**Figure S87:** $\log_2$ Abundance distribution of FGA

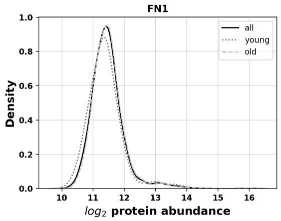


**Figure S88:** $\log_2$ Abundance distribution of FN1

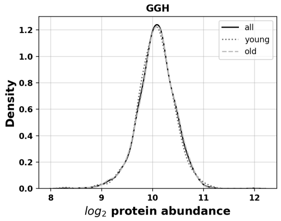


**Figure S89:** $\log_2$ Abundance distribution of GGH

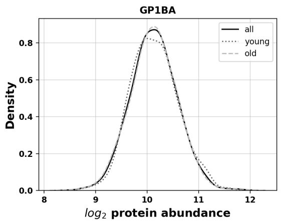


**Figure S90:** $\log_2$ Abundance distribution of GP1BA

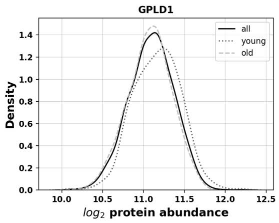


**Figure S91:** $\log_2$ Abundance distribution of GPLD1

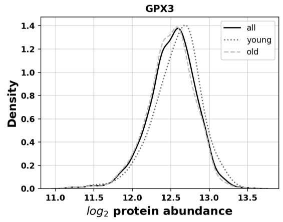


**Figure S92:** $\log_2$ Abundance distribution of GPX3

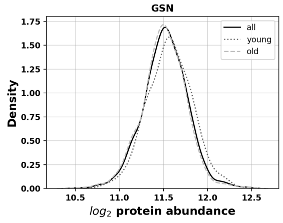


**Figure S93:** $\log_2$ Abundance distribution of GSN

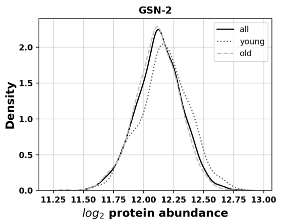


**Figure S94:** $\log_2$ Abundance distribution of GSN-2

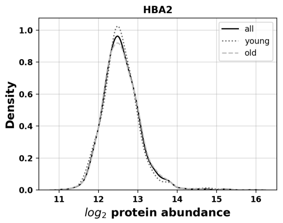


**Figure S95:** $\log_2$ Abundance distribution of HBA2

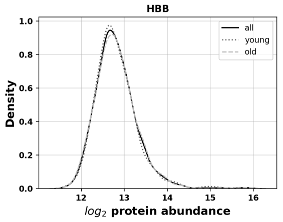


**Figure S96:** $\log_2$ Abundance distribution of HBB

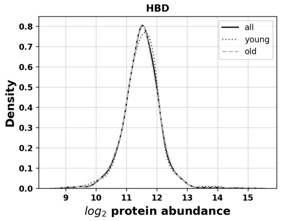


**Figure S97:** $\log_2$ Abundance distribution of HBD

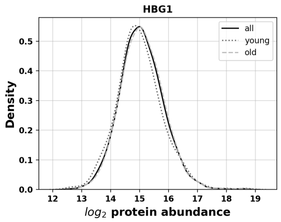


**Figure S98:** $\log_2$ Abundance distribution of HBG1

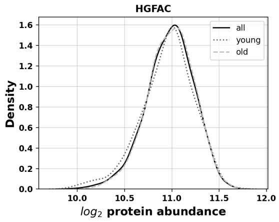


**Figure S99:** $\log_2$ Abundance distribution of HGFAC

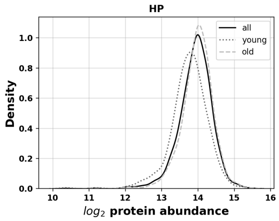


**Figure S100:** $\log_2$ Abundance distribution of HP

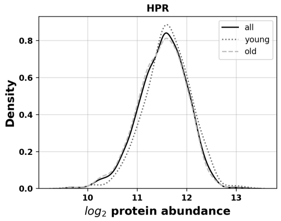


**Figure S101:** $\log_2$ Abundance distribution of HPR

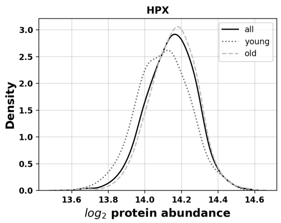


**Figure S102:** $\log_2$ Abundance distribution of HPX

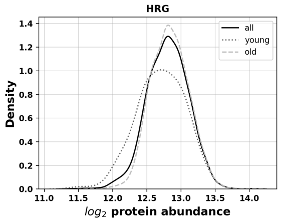


**Figure S103:** $\log_2$ Abundance distribution of HRG

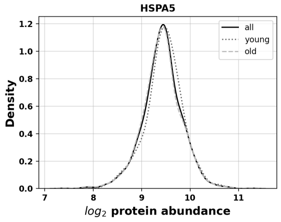


**Figure S104:** $\log_2$ Abundance distribution of HSPA5

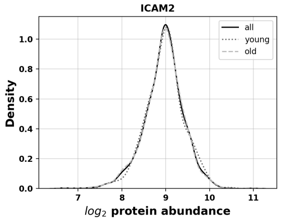


**Figure S105:** $\log_2$ Abundance distribution of ICAM2

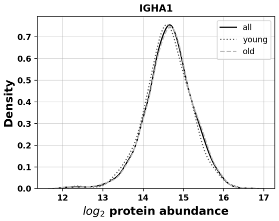


**Figure S106:** $\log_2$ Abundance distribution of IGHA1

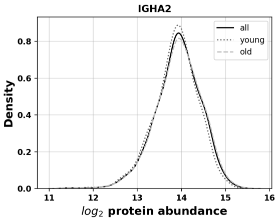


**Figure S107:** $\log_2$ Abundance distribution of IGHA2

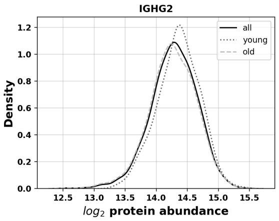


**Figure S108:** $\log_2$ Abundance distribution of IGHG2

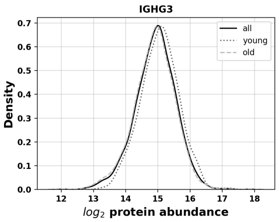


**Figure S109:** $\log_2$ Abundance distribution of IGHG3

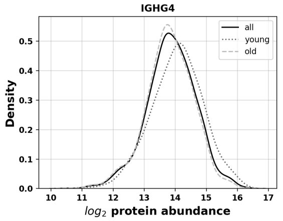


**Figure S110:** $\log_2$ Abundance distribution of IGHG4

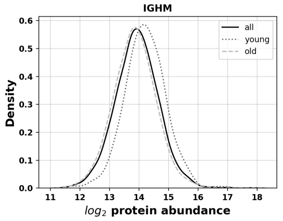


**Figure S111:** $\log_2$ Abundance distribution of IGHM

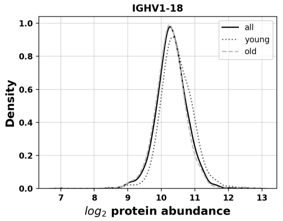


**Figure S112:** $\log_2$ Abundance distribution of IGHV1-18

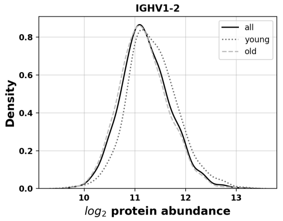


**Figure S113:** $\log_2$ Abundance distribution of IGHV1-2

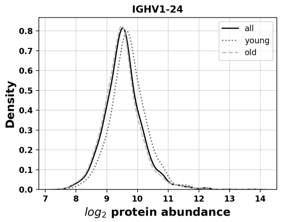


**Figure S114:** $\log_2$ Abundance distribution of IGHV1-24

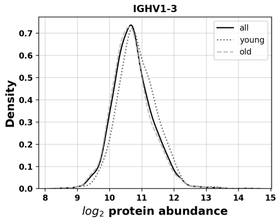


**Figure S115:** $\log_2$ Abundance distribution of IGHV1-3

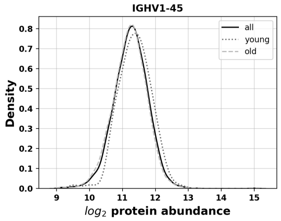


**Figure S116:** $\log_2$ Abundance distribution of IGHV1-45

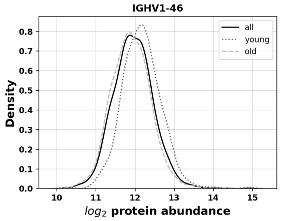


**Figure S117:** $\log_2$ Abundance distribution of IGHV1-46

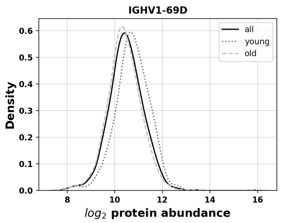


**Figure S118:** $\log_2$ Abundance distribution of IGHV1-69D

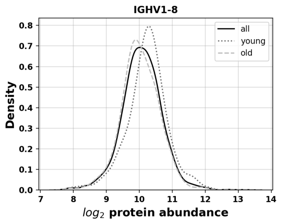


**Figure S119:** $\log_2$ Abundance distribution of IGHV1-8

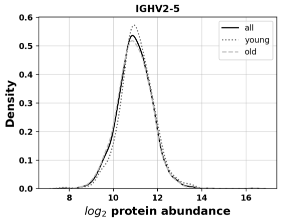


**Figure S120:** $\log_2$ Abundance distribution of IGHV2-5

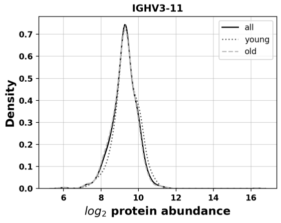


**Figure S121:** $\log_2$ Abundance distribution of IGHV3-11

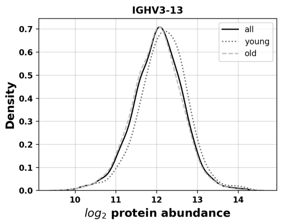


**Figure S122:** $\log_2$ Abundance distribution of IGHV3-13

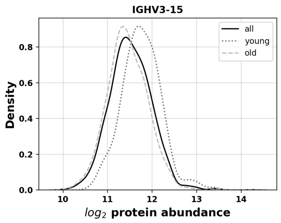


**Figure S123:** $\log_2$ Abundance distribution of IGHV3-15

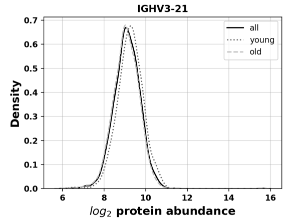


**Figure S124:** $\log_2$ Abundance distribution of IGHV3-21

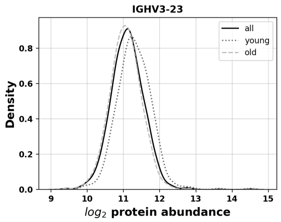


**Figure S125:** $\log_2$ Abundance distribution of IGHV3-23

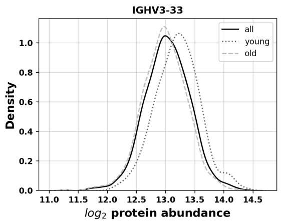


**Figure S126:** $\log_2$ Abundance distribution of IGHV3-33

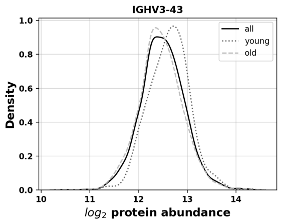


**Figure S127:** $\log_2$ Abundance distribution of IGHV3-43

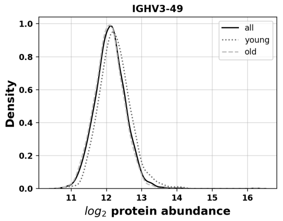


**Figure S128:** $\log_2$ Abundance distribution of IGHV3-49

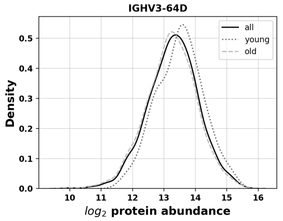


**Figure S129:** $\log_2$ Abundance distribution of IGHV3-64D

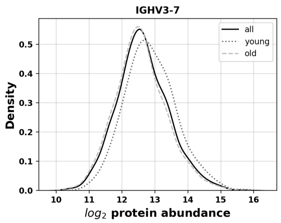


**Figure S130:** $\log_2$ Abundance distribution of IGHV3-7

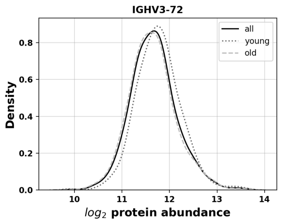


**Figure S131:** $\log_2$ Abundance distribution of IGHV3-72

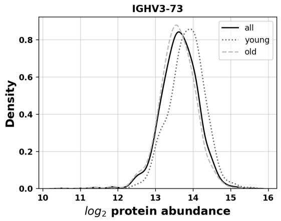


**Figure S132:** $\log_2$ Abundance distribution of IGHV3-73

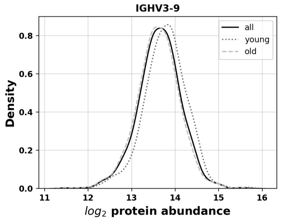


**Figure S133:** $\log_2$ Abundance distribution of IGHV3-9

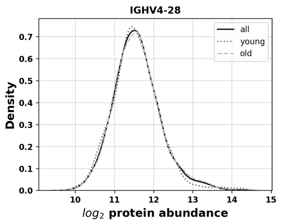


**Figure S134:** $\log_2$ Abundance distribution of IGHV4-28

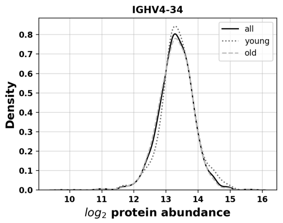


**Figure S135:** $\log_2$ Abundance distribution of IGHV4-34

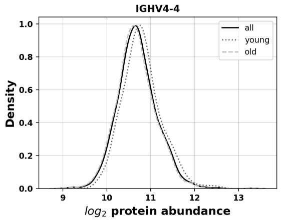


**Figure S136:** $\log_2$ Abundance distribution of IGHV4-4

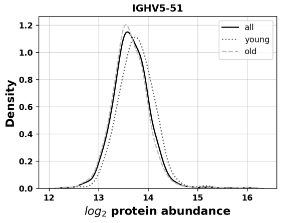


**Figure S137:** $\log_2$ Abundance distribution of IGHV5-51

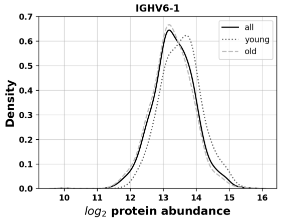


**Figure S138:** $\log_2$ Abundance distribution of IGHV6-1

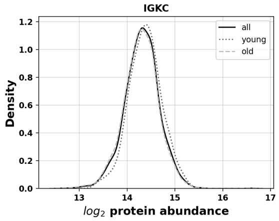


**Figure S139:** $\log_2$ Abundance distribution of IGKC

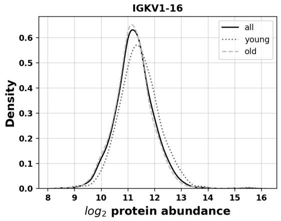


**Figure S140:** $\log_2$ Abundance distribution of IGKV1-16

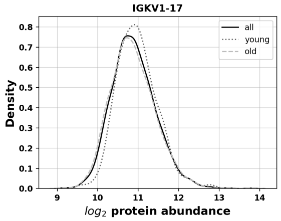


**Figure S141:** $\log_2$ Abundance distribution of IGKV1-17

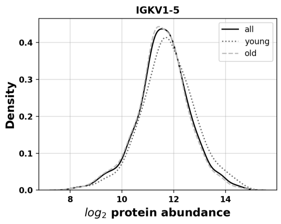


**Figure S142:** $\log_2$ Abundance distribution of IGKV1-5

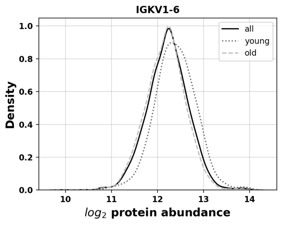


**Figure S143:** $\log_2$ Abundance distribution of IGKV1-6

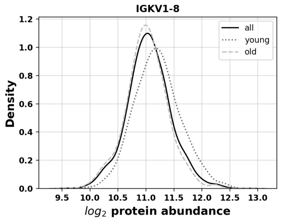


**Figure S144:** $\log_2$ Abundance distribution of IGKV1-8

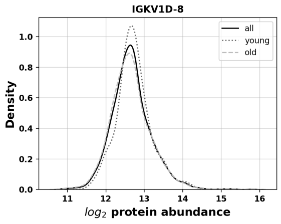


**Figure S145:** $\log_2$ Abundance distribution of IGKV1D-8

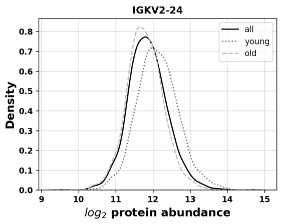


**Figure S146:** $\log_2$ Abundance distribution of IGKV2-24

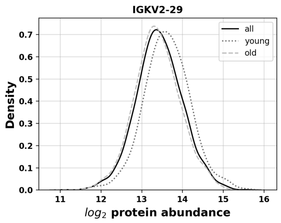


**Figure S147:** $\log_2$ Abundance distribution of IGKV2-29

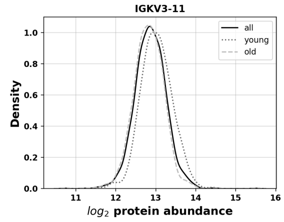


**Figure S148:** $\log_2$ Abundance distribution of IGKV3-11

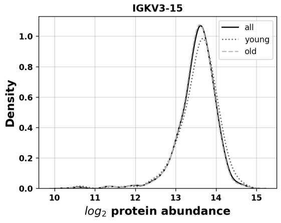


**Figure S149:** $\log_2$ Abundance distribution of IGKV3-15

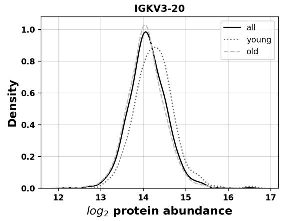


**Figure S150:** $\log_2$ Abundance distribution of IGKV3-20

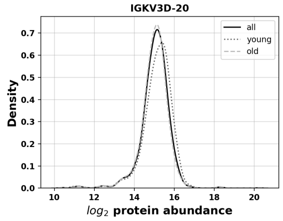


**Figure S151:** $\log_2$ Abundance distribution of IGKV3D-20

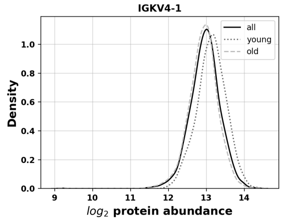


**Figure S152:** $\log_2$ Abundance distribution of IGKV4-1

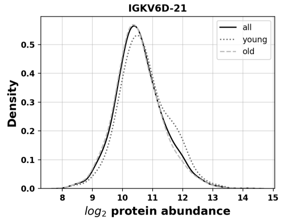


**Figure S153:** $\log_2$ Abundance distribution of IGKV6D-21

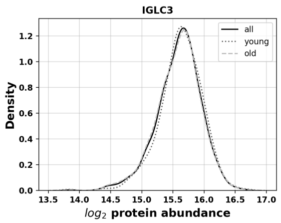


**Figure S154:** $\log_2$ Abundance distribution of IGLC3

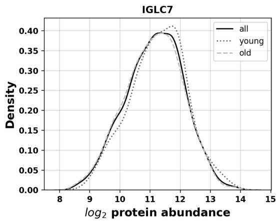


**Figure S155:** $\log_2$ Abundance distribution of IGLC7

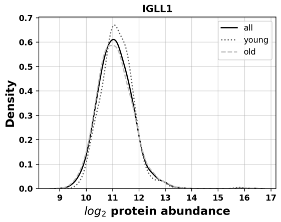


**Figure S156:** $\log_2$ Abundance distribution of IGLL1

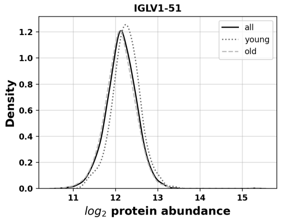


**Figure S157:** $\log_2$ Abundance distribution of IGLV1-51

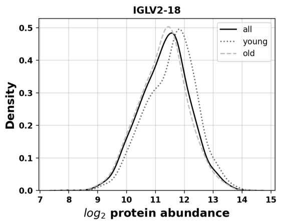


**Figure S158:** $\log_2$ Abundance distribution of IGLV2-18

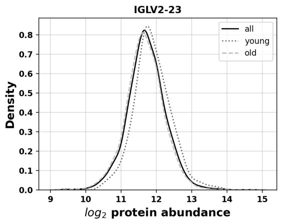


**Figure S159:** $\log_2$ Abundance distribution of IGLV2-23

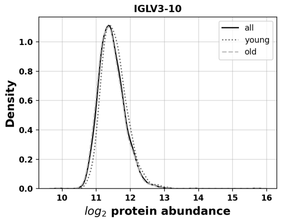


**Figure S160:** $\log_2$ Abundance distribution of IGLV3-10

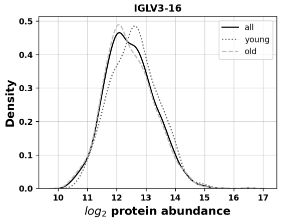


**Figure S161:** $\log_2$ Abundance distribution of IGLV3-16

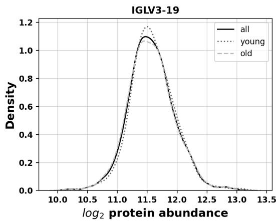


**Figure S162:** $\log_2$ Abundance distribution of IGLV3-19

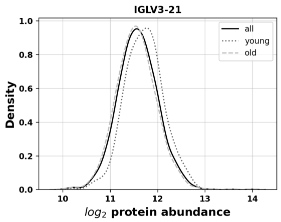


**Figure S163:** $\log_2$ Abundance distribution of IGLV3-21

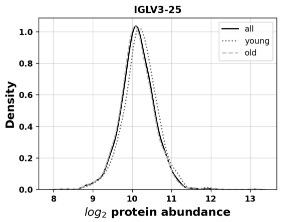


**Figure S164:** $\log_2$ Abundance distribution of IGLV3-25

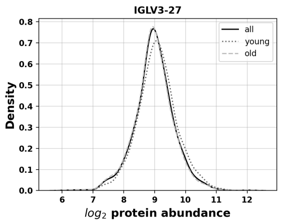


**Figure S165:** $\log_2$ Abundance distribution of IGLV3-27

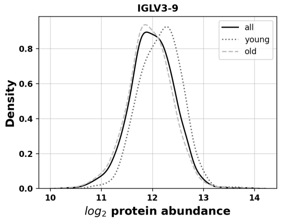


**Figure S166:** $\log_2$ Abundance distribution of IGLV3-9

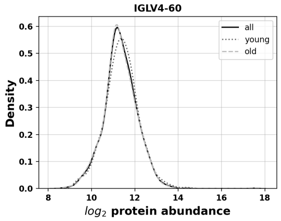


**Figure S167:** $\log_2$ Abundance distribution of IGLV4-60

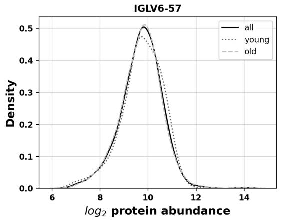


**Figure S168:** $\log_2$ Abundance distribution of IGLV6-57

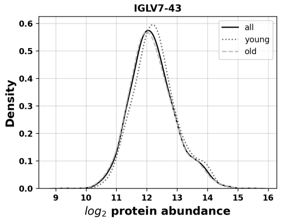


**Figure S169:** $\log_2$ Abundance distribution of IGLV7-43

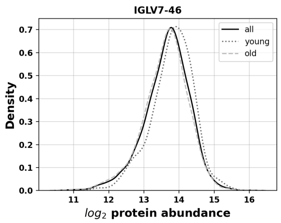


**Figure S170:** $\log_2$ Abundance distribution of IGLV7-46

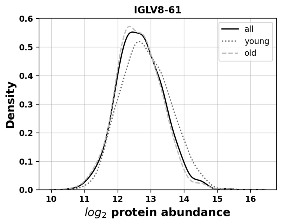


**Figure S171:** $\log_2$ Abundance distribution of IGLV8-61

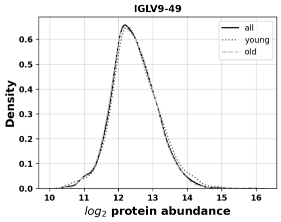


**Figure S172:** $\log_2$ Abundance distribution of IGLV9-49

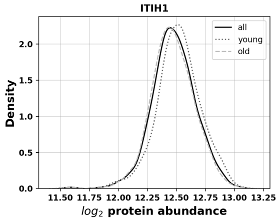


**Figure S173:** $\log_2$ Abundance distribution of ITIH1

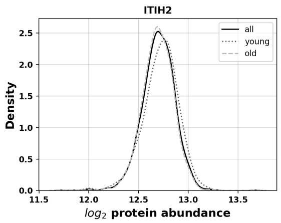


**Figure S174:** $\log_2$ Abundance distribution of ITIH2

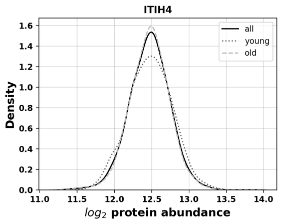


**Figure S175:** $\log_2$ Abundance distribution of ITIH4

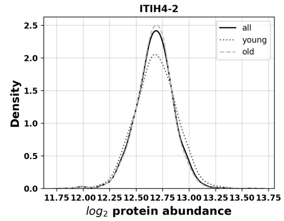


**Figure S176:** $\log_2$ Abundance distribution of ITIH4-2

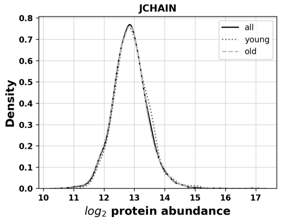


**Figure S177:** $\log_2$ Abundance distribution of JCHAIN

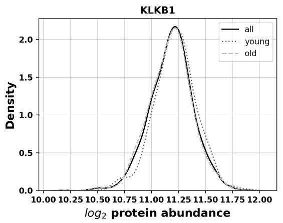


**Figure S178:** $\log_2$ Abundance distribution of KLKB1

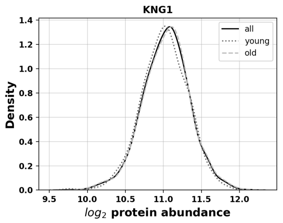


**Figure S179:** $\log_2$ Abundance distribution of KNG1

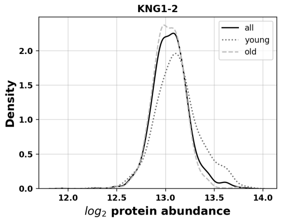


**Figure S180:** $\log_2$ Abundance distribution of KNG1-2

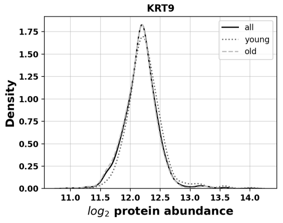


**Figure S181:** $\log_2$ Abundance distribution of KRT9

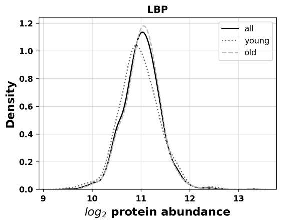


**Figure S182:** $\log_2$ Abundance distribution of LBP

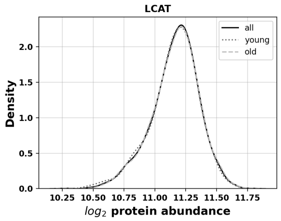


**Figure S183:** $\log_2$ Abundance distribution of LCAT

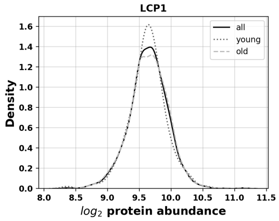


**Figure S184:** $\log_2$ Abundance distribution of LCP1

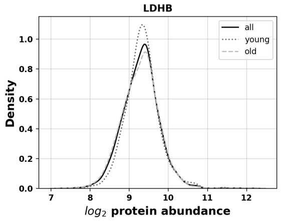


**Figure S185:** $\log_2$ Abundance distribution of LDHB

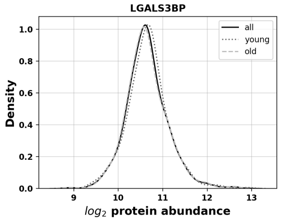


**Figure S186:** $\log_2$ Abundance distribution of LGALS3BP

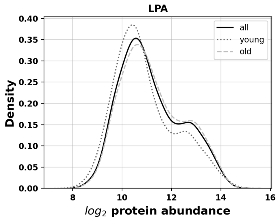


**Figure S187:** $\log_2$ Abundance distribution of LPA

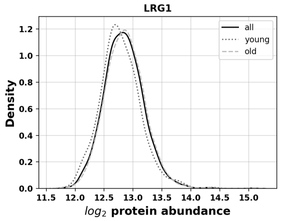


**Figure S188:** $\log_2$ Abundance distribution of LRG1

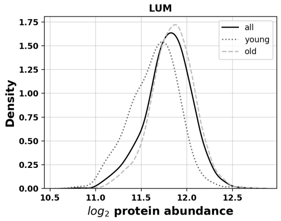


**Figure S189:** $\log_2$ Abundance distribution of LUM

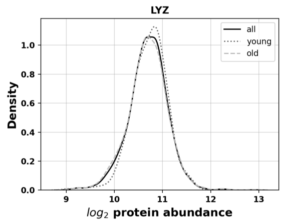


**Figure S190:** $\log_2$ Abundance distribution of LYZ

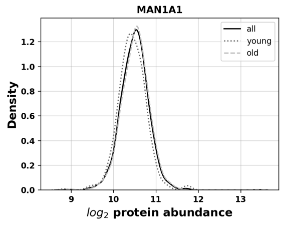


**Figure S191:** $\log_2$ Abundance distribution of MAN1A1

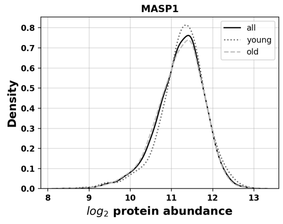


**Figure S192:** $\log_2$ Abundance distribution of MASP1

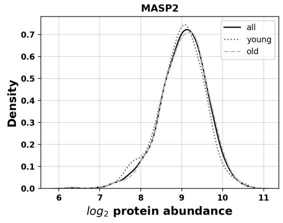


**Figure S193:** $\log_2$ Abundance distribution of MASP2

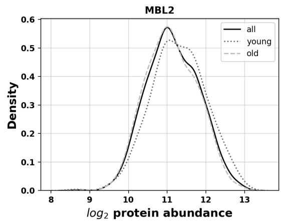


**Figure S194:** $\log_2$ Abundance distribution of MBL2

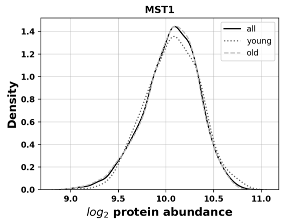


**Figure S195:** $\log_2$ Abundance distribution of MST1

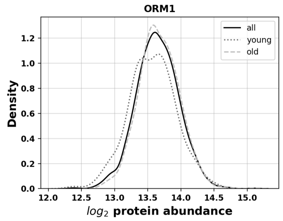


**Figure S196:** $\log_2$ Abundance distribution of ORM1

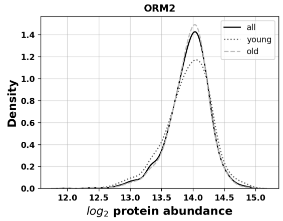


**Figure S197:** $\log_2$ Abundance distribution of ORM2

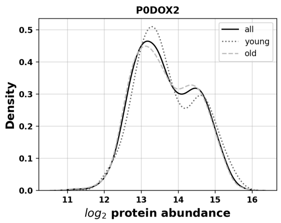


**Figure S198:** $\log_2$ Abundance distribution of P0DOX2

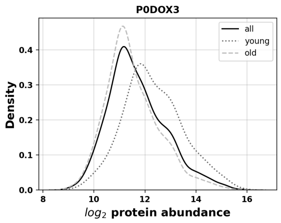


**Figure S199:** $\log_2$ Abundance distribution of P0DOX3

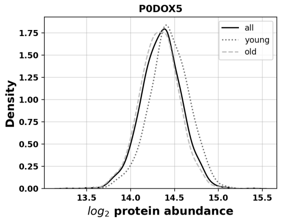


**Figure S200:** $\log_2$ Abundance distribution of P0DOX5

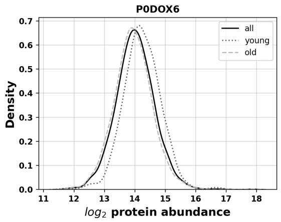


**Figure S201:** $\log_2$ Abundance distribution of P0DOX6

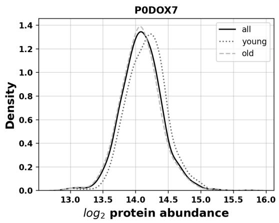


**Figure S202:** $\log_2$ Abundance distribution of P0DOX7

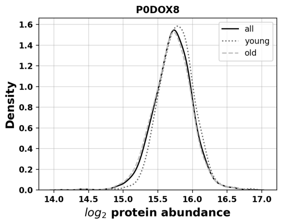


**Figure S203:** $\log_2$ Abundance distribution of P0DOX8

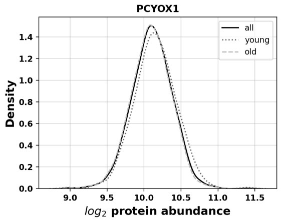


**Figure S204:** $\log_2$ Abundance distribution of PCYOX1

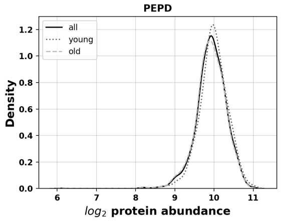


**Figure S205:** $\log_2$ Abundance distribution of PEPD

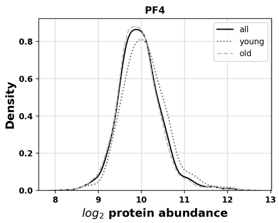


**Figure S206:** $\log_2$ Abundance distribution of PF4

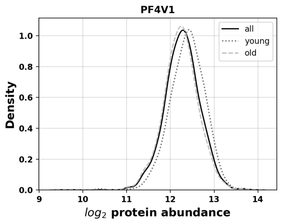


**Figure S207:** $\log_2$ Abundance distribution of PF4V1

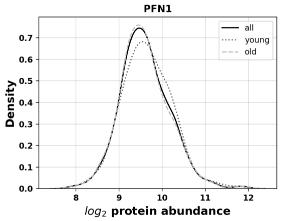


**Figure S208:** $\log_2$ Abundance distribution of PFN1

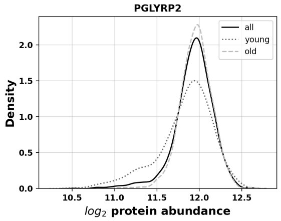


**Figure S209:** $\log_2$ Abundance distribution of PGLYRP2

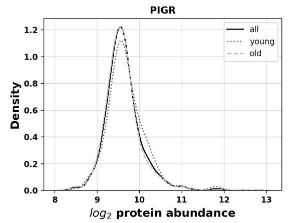


**Figure S210:** $\log_2$ Abundance distribution of PIGR

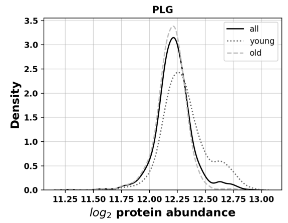


**Figure S211:** $\log_2$ Abundance distribution of PLG

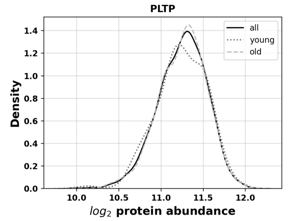


**Figure S212:** $\log_2$ Abundance distribution of PLTP

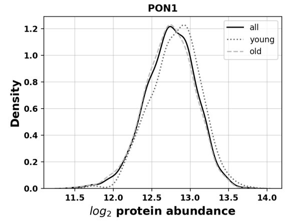


**Figure S213:** $\log_2$ Abundance distribution of PON1

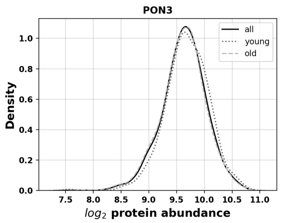


**Figure S214:** $\log_2$ Abundance distribution of PON3

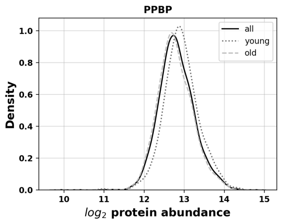


**Figure S215:** $\log_2$ Abundance distribution of PPBP

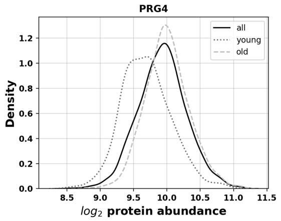


**Figure S216:** $\log_2$ Abundance distribution of PRG4

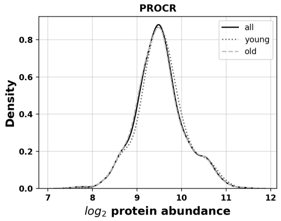


**Figure S217:** $\log_2$ Abundance distribution of PROCR

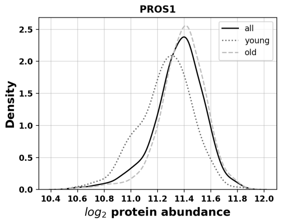


**Figure S218:** $\log_2$ Abundance distribution of PROS1

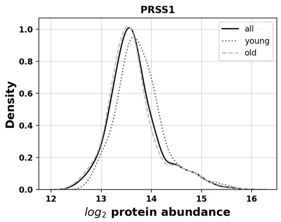


**Figure S219:** $\log_2$ Abundance distribution of PRSS1

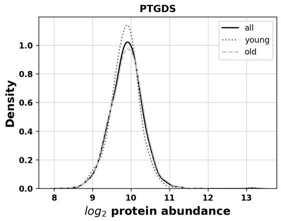


**Figure S220:** $\log_2$ Abundance distribution of PTGDS

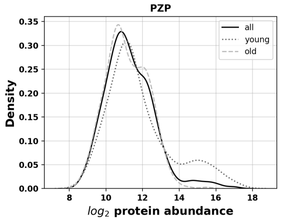


**Figure S221:** $\log_2$ Abundance distribution of PZP

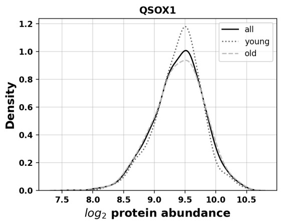


**Figure S222:** $\log_2$ Abundance distribution of QSOX1

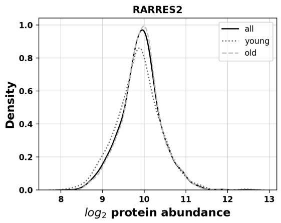


**Figure S223:** $\log_2$ Abundance distribution of RARRES2

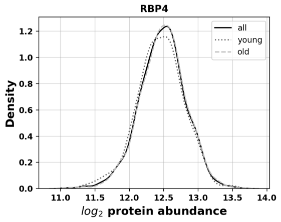


**Figure S224:** $\log_2$ Abundance distribution of RBP4

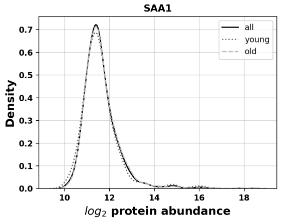


**Figure S225:** $\log_2$ Abundance distribution of SAA1

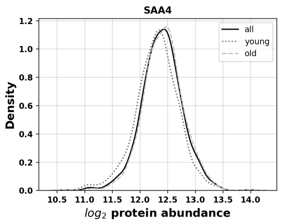


**Figure S226:** $\log_2$ Abundance distribution of SAA4

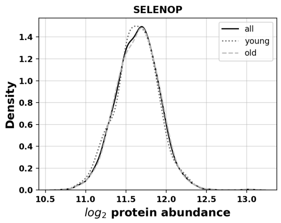


**Figure S227:** $\log_2$ Abundance distribution of SELENOP

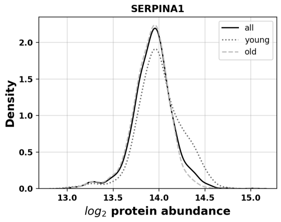


**Figure S228:** $\log_2$ Abundance distribution of SERPINA1

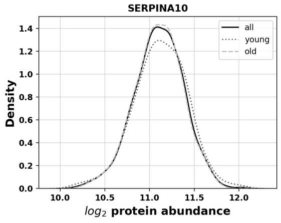


**Figure S229:** $\log_2$ Abundance distribution of SERPINA10

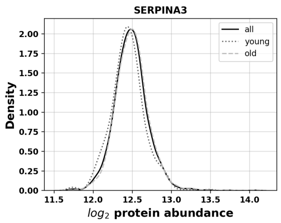


**Figure S230:** $\log_2$ Abundance distribution of SERPINA3

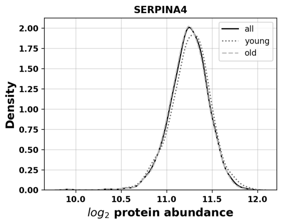


**Figure S231:** $\log_2$ Abundance distribution of SERPINA4

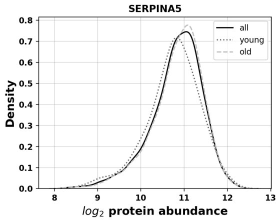


**Figure S232:** $\log_2$ Abundance distribution of SERPINA5

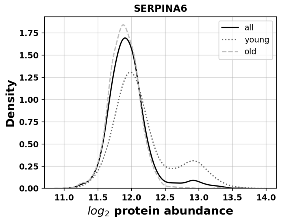


**Figure S233:** $\log_2$ Abundance distribution of SERPINA6

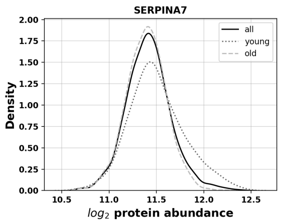


**Figure S234:** $\log_2$ Abundance distribution of SERPINA7

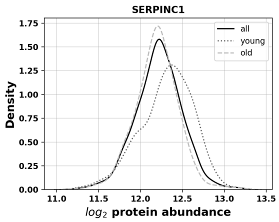


**Figure S235:** $\log_2$ Abundance distribution of SERPINC1

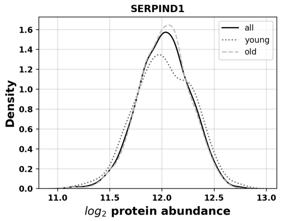


**Figure S236:** $\log_2$ Abundance distribution of SERPIND1

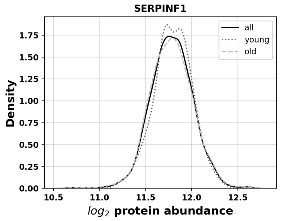


**Figure S237:** $\log_2$ Abundance distribution of SERPINF1

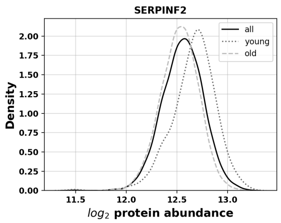


**Figure S238:** $\log_2$ Abundance distribution of SERPINF2

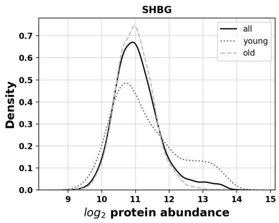


**Figure S239:** $\log_2$ Abundance distribution of SHBG

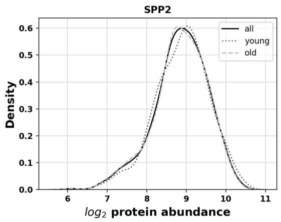


**Figure S240:** $\log_2$ Abundance distribution of SPP2

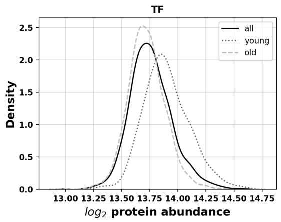


**Figure S241:** $\log_2$ Abundance distribution of TF

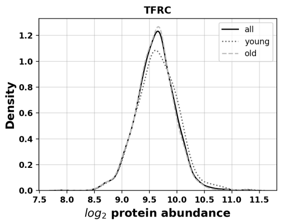


**Figure S242:** $\log_2$ Abundance distribution of TFRC

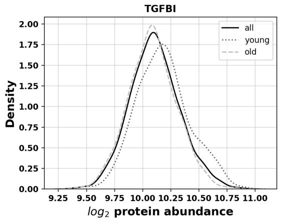


**Figure S243:** $\log_2$ Abundance distribution of TGFBI

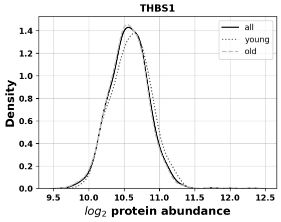


**Figure S244:** $\log_2$ Abundance distribution of THBS1

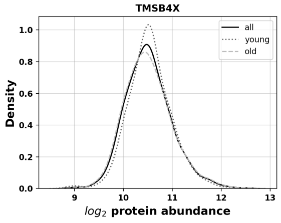


**Figure S245:** $\log_2$ Abundance distribution of TMSB4X

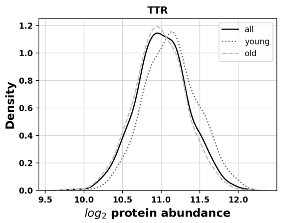


**Figure S246:** $\log_2$ Abundance distribution of TTR

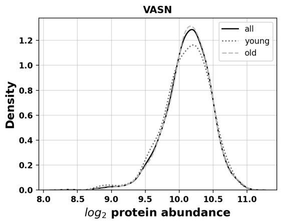


**Figure S247:** $\log_2$ Abundance distribution of VASN

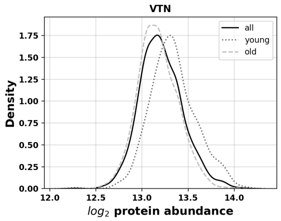


**Figure S248:** $\log_2$ Abundance distribution of VTN

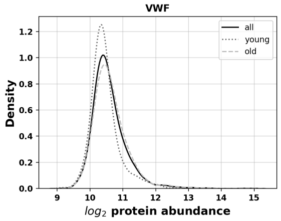


**Figure S249:** $\log_2$ Abundance distribution of VWF

### 2 Correlation between Protein Abundance and Diagnostic Assays

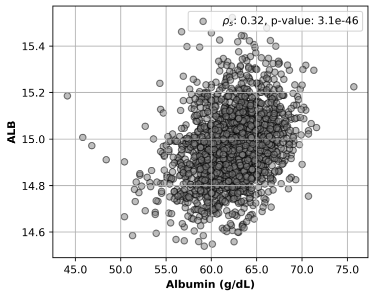


**(a)** Albumin (Diagnostics vs MS)

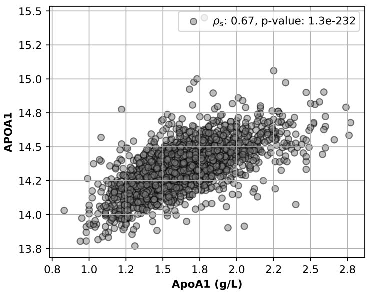


**(b)** Apolipoprotein A1 (Diagnostics vs MS)

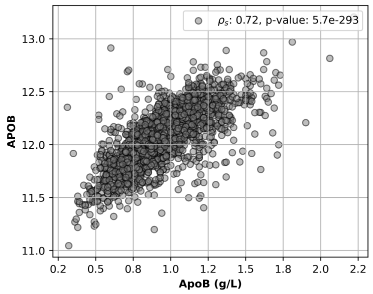


**(c)** Apolipoprotein B (Diagnostics vs MS)

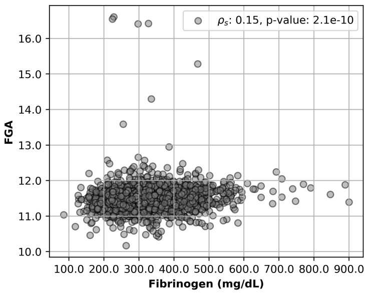


**(d)** Fibrinogen (Diagnostics) vs Fibrinogen Alpha Chain (FGA, MS)

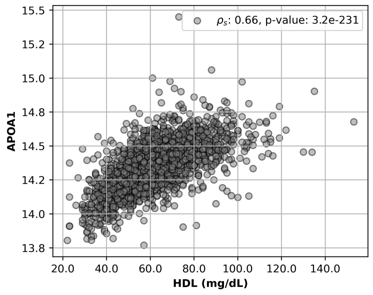


**(e)** High density lipoproteins (HDL, Diagnostics) vs Apolipoprotein A1(APOA1, MS)

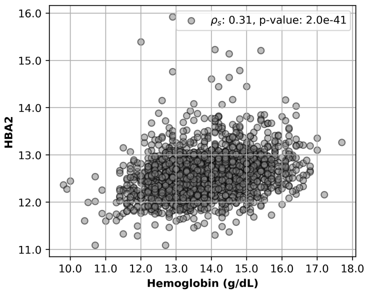


**(f)** Hemoglobin (Diagnostics) vs Hemoglobin Subunit Alpha 2 (HBA2, MS)

**Figure S250:** Scatterplots depicting the relationship between routine labratory measurements and abundance of corresponding proteins.

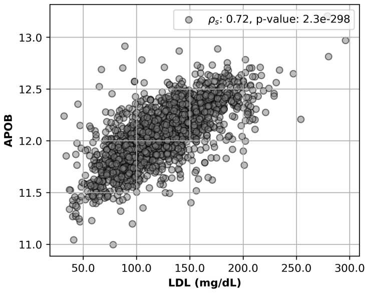


**(g)** Low density lipoproteins (LDL, Diagnostics) vs Apolipoprotein B (APOB, MS)

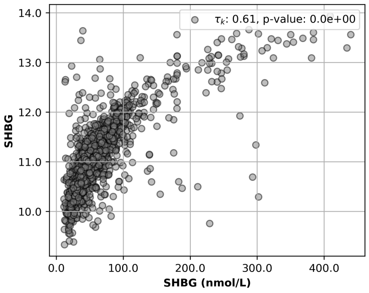


**(h)** Sex hormone binding globuline (Diagnostics) vs Sex hormone binding globuline (SHBG, MS)

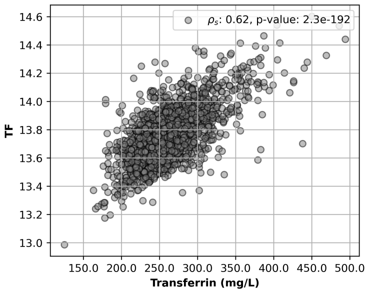


**(i)** Transferrin (Diagnostics) vs Transferrin (TF, MS)

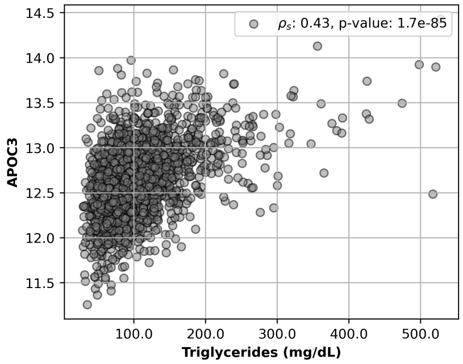


**(j)** Tryglycerides (Diagnostics) vs Apolipoprotein C3 (APOC3, MS)

**Figure S250:** Scatterplots depicting the relationship between routine laboratory measurements and abundance of corresponding proteins.

### 3 Additional Excel Tables

**Table S2: Results of functional analysis with PANTHER classification system**
**Table S3: Results of association analysis for the whole cohort**
**Table S4: Results of association analysis for the older age group**
**Table S5: Results of association analysis for the younger age group**

### 4 Effect Size Concurrence between CHRIS and BASE II

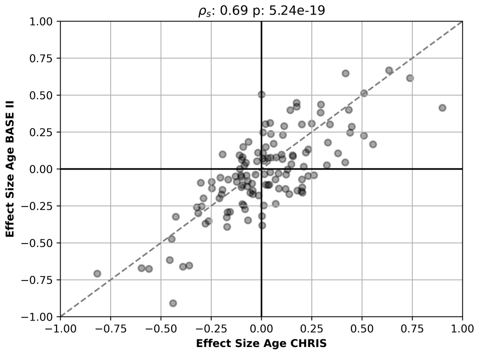


**(k)** Age

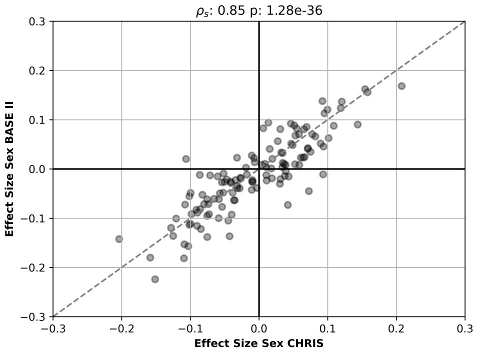


**(l)** Sex

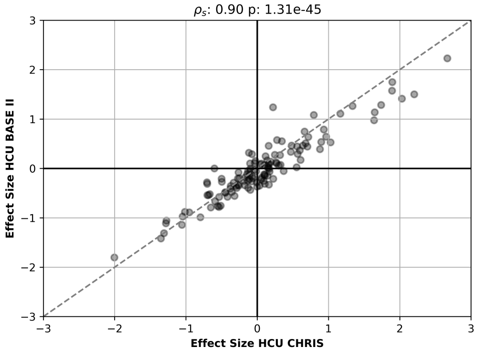


**(m)** HCU

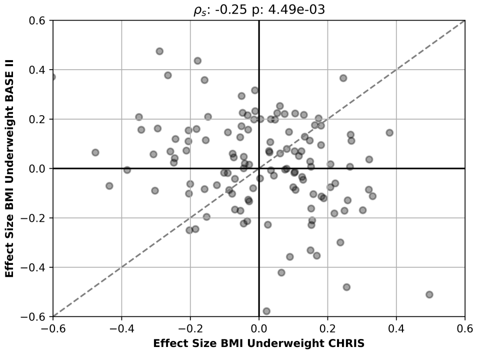


**(n)** BMI Underweight

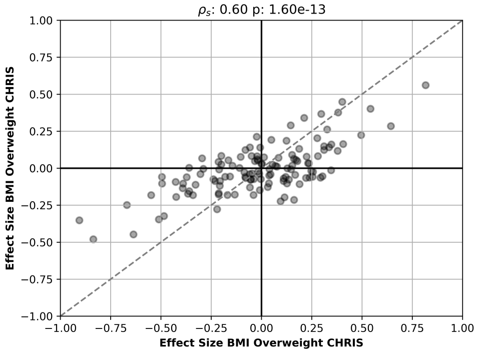


**(o)** BMI Overweight

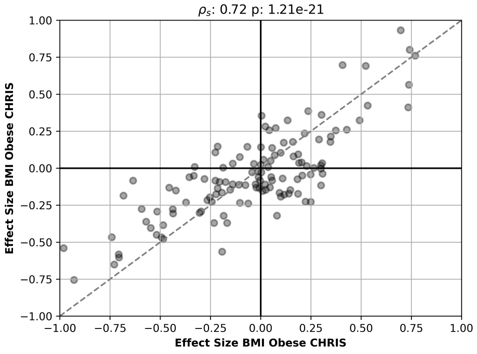


**(p)** BMI Obese

**Figure S251:** Scatterplots depicting the concordance of effect sizes among the proteins present in the proteomic datasets of BASE II and CHRIS[1]. Correlation coefficients are calculated with Spearman's rho. Dashed lines represent the identity line.

### 5 Influence of MHT on the Serum Proteome of BASE II Participants

**Table S6:** Proteins significantly associated with ongoing MHT in women of the older age group and corresponding adjusted p-values, coefficients and effect sizes.

| Gene Symbol | Uniprot ID | Protein Name | $p_{adj}$ | $coef$ | $es$ |
|---|---|---|---|---|---|
| AGT | P01019 | angiotensinogen | 1.25e-04 | 0.24 | 0.89 |
| PLG | P00747 | plasminogen | 3.41e-02 | 0.10 | 0.68 |
| PGLYRP2 | Q96PD5 | peptidoglycan recognition protein 2 | 3.91e-02 | -0.12 | -0.67 |

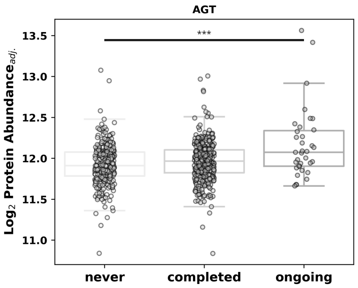


**(a)** AGT

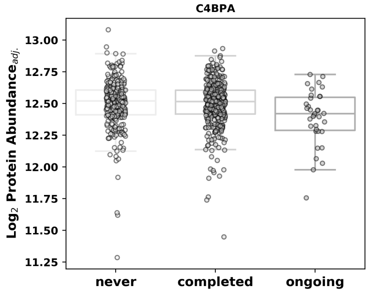


**(b)** C4BPA

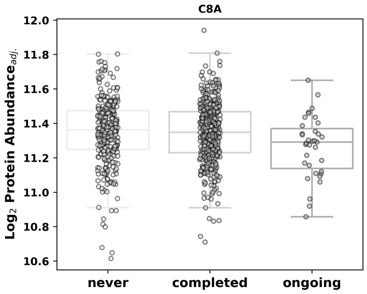


**(c)** C8

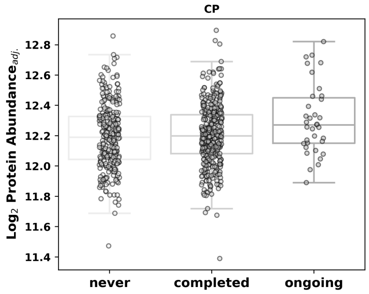


**(d)** CP

**Figure S252:** Boxplots showing the distributions of selected proteins (adjusted by age and BMI) for BASE II study participants, who never underwent MHT, of those who completed MHT and those who are currently under treatment. Asteriks indicating the unadjusted significance of Mann-Whitney-U tests. (* is indicating a p-value <0.05, ** <0.01, *** <0.001)

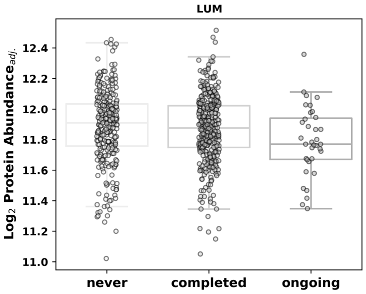


**(e)** LUM

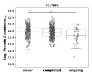

**(f)** PG-LYRP2

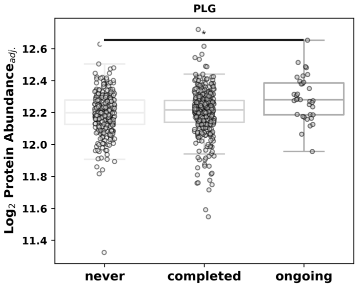


**(g)** PLG

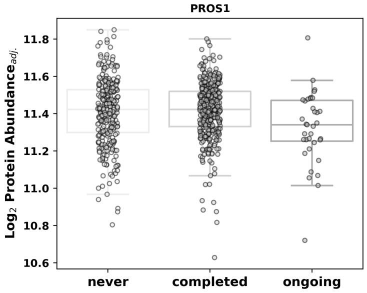


**(h)** PROS1

**Figure S252:** Boxplots showing the distributions of selected proteins (adjusted by age and BMI) for BASE II study participants, who never underwent MHT, of those who completed MHT and those who are currently under treatment. Asteriks indicating the unadjusted significance of Mann-Whitney-U tests. (* is indicating a p-value <0.05, ** <0.01, *** <0.001)

**Table S7:** Median and IQR values for age and sex demographics of matched groups (Control, Treatment) used for classification of MHT

| Covariates | Control Group | MHT Treatment Group |
|---|---|---|
| N | 35 | 35 |
| Age (in years) | 68.6 [4.6] | 68.7 [4.3] |
| BMI | 25.2 [5.7] | 25 [5.6] |

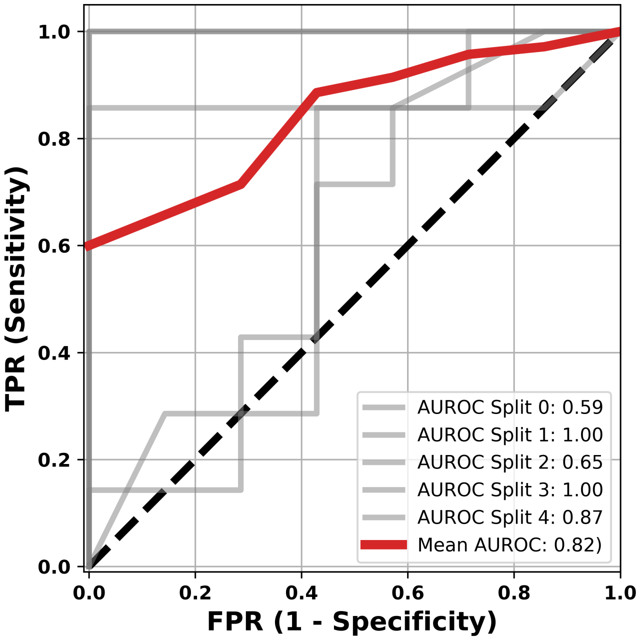


**Figure S253:** Receiver Operating Characteristic (ROC) curve illustrating the performance of different splits and its mean during the CV process of the random forest model, in classifying ongoing Menopausal Hormone Therapy (MHT).

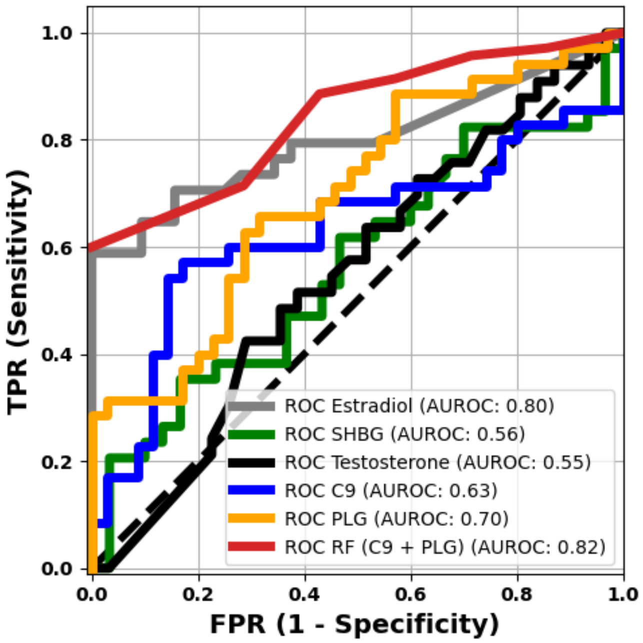


**Figure S254:** Receiver Operating Characteristic (ROC) curve illustrating the performance of CP, PLG, SHBG, and Testosterone levels, along with the trained Random Forest model, in classifying ongoing Menopausal Hormone Therapy (MHT).

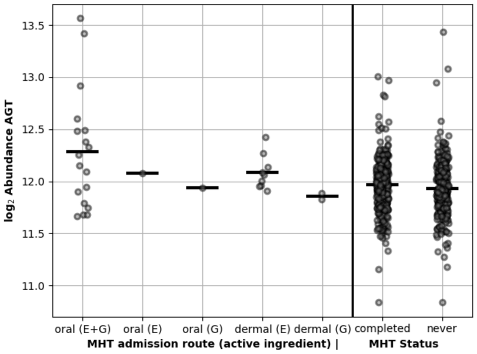


**(a)** AGT

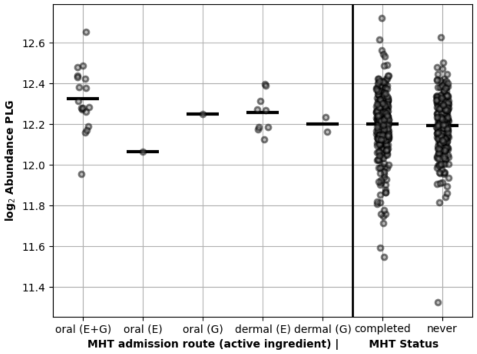


**(b)** PLG

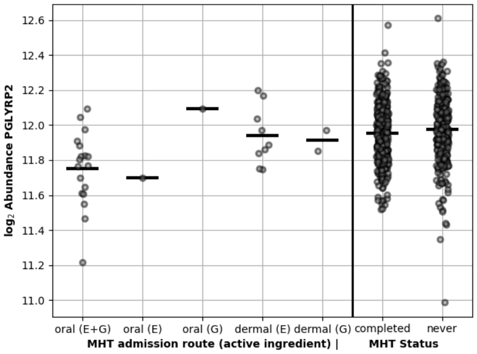


**(c)** PGLYRP2

**Figure S255:** Scatter plot showing log2 abundance of proteins associated with MHT of BASE II participants with ongoing MHT stratified by admission routes and active ingridients (E = Estrogen, G = Gestagen) and different admission routes. Right of the vertical line, log2 protein abundance of women are display that are not under MHT treatment at sampling time.

### 6 Association Analysis for Lifestyle Factors and Comorbidities

**Table S8:** Proteins significantly associated with metabolic syndrome in the whole cohort and corresponding adjusted p-values, coefficients and effect sizes.

| Gene Symbol | Uniprot ID | Protein Name | $p_{adj}$ | $coef$ | $es$ |
|---|---|---|---|---|---|
| APOB | P04114 | apolipoprotein B | 2.20e-04 | 0.08 | 0.27 |
| APOD | P05090 | apolipoprotein D | 1.14e-09 | -0.12 | -0.37 |
| APOC4 | P55056 | apolipoprotein C4 | 4.53e-12 | 0.18 | 0.44 |
| APOC2 | P02655 | apolipoprotein C2 | 2.45e-14 | 0.21 | 0.46 |

**Table S9:** Proteins significantly associated with diabetes in the old cohort and corresponding adjusted p-values, coefficients and effect sizes.

| Gene Symbol | Uniprot ID | Protein Name | $p_{adj}$ | $coef$ | $es$ |
|---|---|---|---|---|---|
| APOA4 | P06727 | apolipoprotein A4 | 8.99e-04 | 0.13 | 0.38 |
| APOD | P05090 | apolipoprotein D | 3.15e-03 | -0.11 | -0.34 |

**Table S10:** Proteins significantly associated with diabetes when accounting for metabolic syndrome in the old cohort and corresponding adjusted p-values, coefficients and effect sizes.

| Gene Symbol | Uniprot ID | Protein Name | $p_{adj}$ | $coef$ | $es$ |
|---|---|---|---|---|---|
| APOA4 | P06727 | apolipoprotein A4 | 9.63e-06 | 0.17 | 0.49 |

**Table S11:** Proteins significantly associated with current smoking in the whole cohort and corresponding adjusted p-values, coefficients and effect sizes.

| Gene Symbol | Uniprot ID | Protein Name | $p_{adj}$ | $coef$ | $es$ |
|---|---|---|---|---|---|
| IGHG2 | P01859 | immunoglobulin heavy constant gamma 2 | 2.11e-04 | -0.18 | -0.46 |
| PIGR | P01833 | polymeric immunoglobulin receptor | 4.49e-12 | 0.31 | 0.71 |

**Table S12:** Proteins significantly associated with excessive drinking (> 30 g per day) in the whole cohort and corresponding adjusted p-values, coefficients and effect sizes.

| Gene Symbol | Uniprot ID | Protein Name | $p_{adj}$ | $coef$ | $es$ |
|---|---|---|---|---|---|
| C9 | P02748 | complement C9 | 1.47e-03 | -0.13 | -0.42 |
| CNDP1 | Q96KN2 | carnosine dipeptidase 1 | 7.36e-08 | 0.16 | 0.60 |
| SERPINA7 | P05543 | serpin family A member 7 | 2.85e-08 | -0.13 | -0.57 |
| F12 | P00748 | coagulation factor XII | 4.92e-02 | 0.12 | 0.35 |
| APOC3 | P02656 | apolipoprotein C3 | 4.60e-10 | 0.27 | 0.67 |
| RBP4 | P02753 | retinol binding protein 4 | 9.84e-07 | 0.19 | 0.56 |
| APOC2 | P02655 | apolipoprotein C2 | 1.32e-02 | 0.17 | 0.37 |
| HPR | P00739 | haptoglobin-related protein | 2.63e-02 | 0.18 | 0.37 |
